# Learning the fitness dynamics of pathogens from phylogenies

**DOI:** 10.1101/2023.12.23.23300456

**Authors:** Noémie Lefrancq, Loréna Duret, Valérie Bouchez, Sylvain Brisse, Julian Parkhill, Henrik Salje

**Affiliations:** Department of Genetics, University of Cambridge, Cambridge, UK; Department of Veterinary Medicine, University of Cambridge, Cambridge, UK; Institut Pasteur, Université de Paris, Biodiversity and Epidemiology of Bacterial Pathogens, Paris, France; National Reference Center for Whooping Cough and Other Bordetella Infections, Paris, France

## Abstract

The dynamics of pathogen genetic diversity, including the emergence of lineages with increased fitness, is a foundational concept of disease ecology with key public health implications. However, the identification of distinct lineages and estimation of associated fitness remain challenging, and are rarely done outside densely sampled systems. Here, we present a scalable framework that summarizes changes in population composition in phylogenies, allowing for the automatic detection of lineages based on shared fitness and evolutionary relationships. We apply our approach to a broad set of viruses and bacteria (SARS-CoV-2, H3N2 influenza, *Bordetella pertussis* and *Mycobacterium tuberculosis)* and identify previously undiscovered lineages, as well as specific amino acid changes linked to fitness changes, the findings of which are robust to uneven and limited observation. This widely-applicable framework provides an avenue to monitor evolution in real-time to support public health action and explore fundamental drivers of pathogen fitness.

**One sentence summary:** Using an agnostic framework we shed light on changes in population composition in phylogenetic trees, allowing for the automatic detection of circulating lineages and estimation of fitness dynamics.

## Main text

For most pathogens, there are constantly changing patterns of strain composition. Pressures to evade host immunity, environmental shifts or changing abilities to infect and disseminate in hosts result in the emergence of some lineages and the extinction of others. These dynamic patterns of genetic diversity are a fundamental aspect of disease ecology. They also have potentially critical public health implications, including signifying immune or vaccine escape or improved transmissibility. It has, however, been difficult to identify and quantify lineages with differential levels of fitness, especially outside highly genetically sampled pathogen systems such as SARS-CoV-2 or influenza(*1–3*). Identifying lineages with improved fitness would allow focused public health response, through e.g., targeted vaccination, as well as provide key insights into the underlying ecology of disease systems.

Existing methods to monitor the fitness of strains at the population level mostly rely on *a priori* lineage definitions, for example, Pango lineages(*4*) or Nextstrain Clades(*5*), the global clades for influenza(*6*), or strains defined by pre-determined single mutations for *Bordetella pertussis(7*). Strain fitness can be estimated using models that capture the changing proportion of individual lineages through time, typically with multinomial logistic models. These models are computationally efficient and provide key insights, for example, to track the effect of amino-acid substitutions(*8*), or vaccine implementation(*3, 9*) on fitness. However, these approaches rely on an ability to group individual sequences into different lineages, which is usually based on consensus opinion, arbitrary thresholds in amino acid difference and importantly, unlinked to underlying differences in fitness. This is problematic as it means we are not reliably capturing emergent lineages with increased fitness.

Phylogenetic tree-based methods provide an alternative strategy to uncover strain fitness. Strains with increased fitness will transmit more frequently, leading to a higher branching rate in the phylogeny and more sampled descendants. The fitness of lineages can therefore be inferred from their branching pattern in a phylogeny using phylodynamic approaches such as birth-death models(*10*). Multi-type birth-death models extend this idea by allowing the birth and death rate of lineages, and thereby fitness, to depend on a lineage’s state or type, which may be known (e.g. genotype, mutations(*11, 12*)) or inferred(*13*). However, these models are computationally challenging to run, especially given the large amount of data now being generated. They are also susceptible to sampling biases in both space and time, which are common in phylogenetic analyses. There are alternative approaches that focus on the broad population structure(*14*) or changes in effective population size(*15*) but are not able to capture lineage fitness. Other works(*2, 10, 16*) have been done at a more granular level, but do not allow for a broad understanding of fitness changes through time.

Here we present a novel agnostic framework that summarizes the changes in population composition in phylogenetic trees through time, allowing for the automatic detection of circulating lineages based on differences in fitness, which we quantify and link back to specific amino acid changes. We apply this approach to SARS-CoV-2, influenza H3N2, *Bordetella pertussis (B. pertussis)* and *Mycobacterium tuberculosis (M. tuberculosis)*. We selected these respiratory pathogens as they present a diverse set of viruses and bacteria at both local and global scales, and include both well-studied and understudied threats to human health. Taking each pathogen in turn, we use our novel analytical framework to make critical insights into the set of discrete lineages circulating over time, their individual fitness, as well as the genomic changes linked to quantified shifts in fitness.

### Tracking population composition in timed phylogenetic trees

Our framework builds on a genetic distance-based index that measures the epidemic success of each node (internal or terminal) in a time-resolved phylogeny (Figure 1A)(*16*). This measure is based on the expectation that nodes sampled from an emerging fitter lineage will be phylogenetically closer than the rest of the population at that time. The index of each node is derived from the distance distribution from that node to all other nodes that circulate at that time, weighted by a kernel with a set timescale. This weight allows us to track lineage emergence dynamically, focusing on short distances between nodes (containing information about recent population dynamics) rather than long distances (containing information about past evolution). The timescale is tailored to the specific pathogen studied and its choice will depend on the molecular signal, as well as the transmission rate. Using the principles of coalescent theory in structured populations(*17–19*), we derive the expected index dynamics through time in the case of an emerging successful lineage (Figure 1A, derivation in Supplementary Material). The dynamics of this index summarize changes in the composition of populations over time, linked to fitness at the population level (Figure 1B-E and S1).

**Figure 1:**
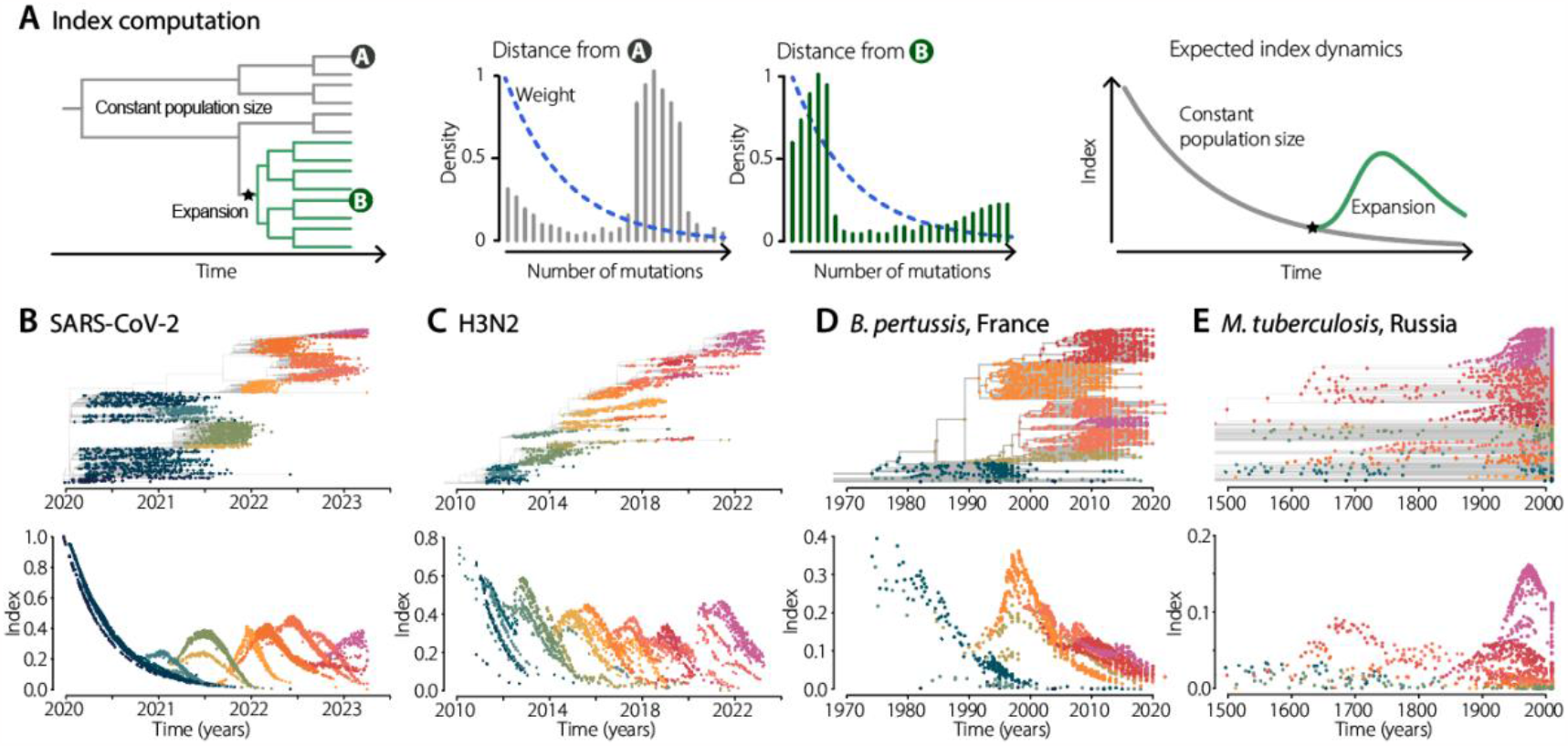
Tracking changes in population composition by following index dynamics. **(A)** Schematics describing the principles of index computation. From left to right: example of a time-resolved phylogenetic tree with a background population (gray) and an emerging lineage (green); pairwise distance distribution from terminal node A, or terminal node B, respectively, to the rest of the population, with the dashed blue line denoting the geometric weighting; and expected index dynamics over time. See methods for details. **(B-D)** For each pathogen, we present the index dynamics computed at each node (terminal or internal). Colors represent the different lineages identified by their different index dynamics (Figure S2). Dynamics colored by known lineages are presented in Figure S1.

### Agnostic identification of pathogen lineages

We developed a tree partitioning algorithm using generalized additive models that finds the set of lineages that best explains the index dynamics (Figure 1B-E and S2). We assessed the generality of our approach across viruses and bacteria by analyzing four pathogens: SARS-CoV-2 (N=3129 global whole genome sequences), influenza H3N2 (N=1476 global hemagglutinin [HA] sequences), *B. pertussis* (N=1248 whole genome sequences from France) and *M. tuberculosis* (N=998 whole genome sequences from Samara, Russia(*20*)). All four are respiratory pathogens whose spread and fitness have been previously studied using genomic data. We found that our framework was able to capture the lineage dynamics of each pathogen considered (Figure 1B-E). Using this framework on SARS-CoV-2 worldwide, we agnostically tracked the changes in population composition (Figure 1B), with each main variant of concern having a clear change in index dynamics. Further, we found that our framework was able to capture population changes for the variety of pathogens considered. Clade replacement was tracked in the influenza H3N2 time-resolved worldwide phylogeny (Figure 1C), despite the gene marker length being small (1698 bp). Going beyond RNA viruses, we tested our model on two bacteria, *B. pertussis* in France and *M. tuberculosis* in Samara, Russia, with largely different diversity and time scales (Figure 1D-E). In both cases, our framework was able to track changes in the population composition, allowing us to refine the *a priori-*defined lineages. Our framework provides an insightful summary of the changes in population structure, by only following the index dynamics.

### Pathogen lineages agnostically identified in the context of previous studies

For each pathogen, we explored how our automatic classification relates to previously identified lineages (Figure 2). We computed the Adjusted Rand-Index (ARI) to measure the agreement between classifications, accounting for random clustering(*21*). A value of 1 corresponds to perfect agreement with previously identified lineages, whereas a value of 0 would be expected if clusters were assigned at random. Overall, we found that our agnostic identification of lineages was in agreement with current classifications (mean ARI of 0.75 across pathogens, min 0.62, max 0.94). The five SARS-CoV-2 variants of concern that spread globally were perfectly delineated by our framework (Alpha [B.1.1.7; 20I], Beta [B.1.351; 20H], Gamma [P.1.*; 20J], Delta [B.1.617.2/AY.*; 21A/21J], and Omicron [BA.1.1.529/BA.*; 21K])(*22, 23*), and the majority of sub-variants were correctly called as well (ARI = 0.80, Figure 2A). We noted that sub-variants that reached a maximum proportion of less than 5% in our global dataset were indistinguishable from others. This highlights the power of our framework in finding lineages that emerge at the geographical scale of the dataset, i.e. globally. Replicating the analysis to SARS-CoV-2 datasets by continent, we re-identify the variants of interest that mainly spread within those continents, e.g. Eta/B.1.525 in Africa, Mu/B.1.621 in the Americas and EU1 in Europe (Figure S3-4)(*24*– *26*). We found similar results for H3N2, with the global subclades being well-matched (ARI of 0.62). Our agnostic framework mainly differed from the existing classification when considering global clades at a very low frequency in our dataset (for example clades 1*, only 2% of sequences). This further highlights that our framework is focusing on the broad population changes. *B. pertussis*’s population composition is less well-studied. To date, only a few clades have been reported, defined by changes in alleles of the promoter of the pertussis toxin (ptxP) and fimbriae 3 gene (fim3)(*7*). Our framework was able to find these major clades (ARI = 0.63). We further found three new lineages that emerged. These three lineages have clear distinct index dynamics (Figure 1D, pink, red and purple lineages), but have not been previously identified. Further, we recovered most of the known *M. tuberculosis* lineages and sublineages (ARI = 0.92). Specifically, the main global lineages were found(*20, 27, 28*), with the exception of the distinction between the Central Asian Strain (CAS) and East African Indian (EAI) lineages, which are both present in very small numbers in the dataset and therefore indistinguishable. The SNP-defined sub-lineages were mostly recovered(*29*), with some discrepancy in lineages such as Harleem, Ural and Latin American-Mediterranean (LAM), which can be attributed to the index focusing on signal of lineage expansion rather than a SNP definition. Therefore, our analysis was able to track the expansion of those lineages specifically in Samara, Russia, rather than the global sub-lineages that might have first expanded elsewhere. This highlights the granularity of our framework, which is able to track lineage expansion at a local level. To investigate how our framework compares to existing ones, we compared the (sub-)lineages in SARS-CoV-2, the pathogen system with the most well-characterized lineages, from our approach with that identified using fastbaps(*14*) and treestructure(*15*). We found that by specifically considering the fitness of the lineages, we could more consistently recover the known lineages (Figure S5).

**Figure 2:**
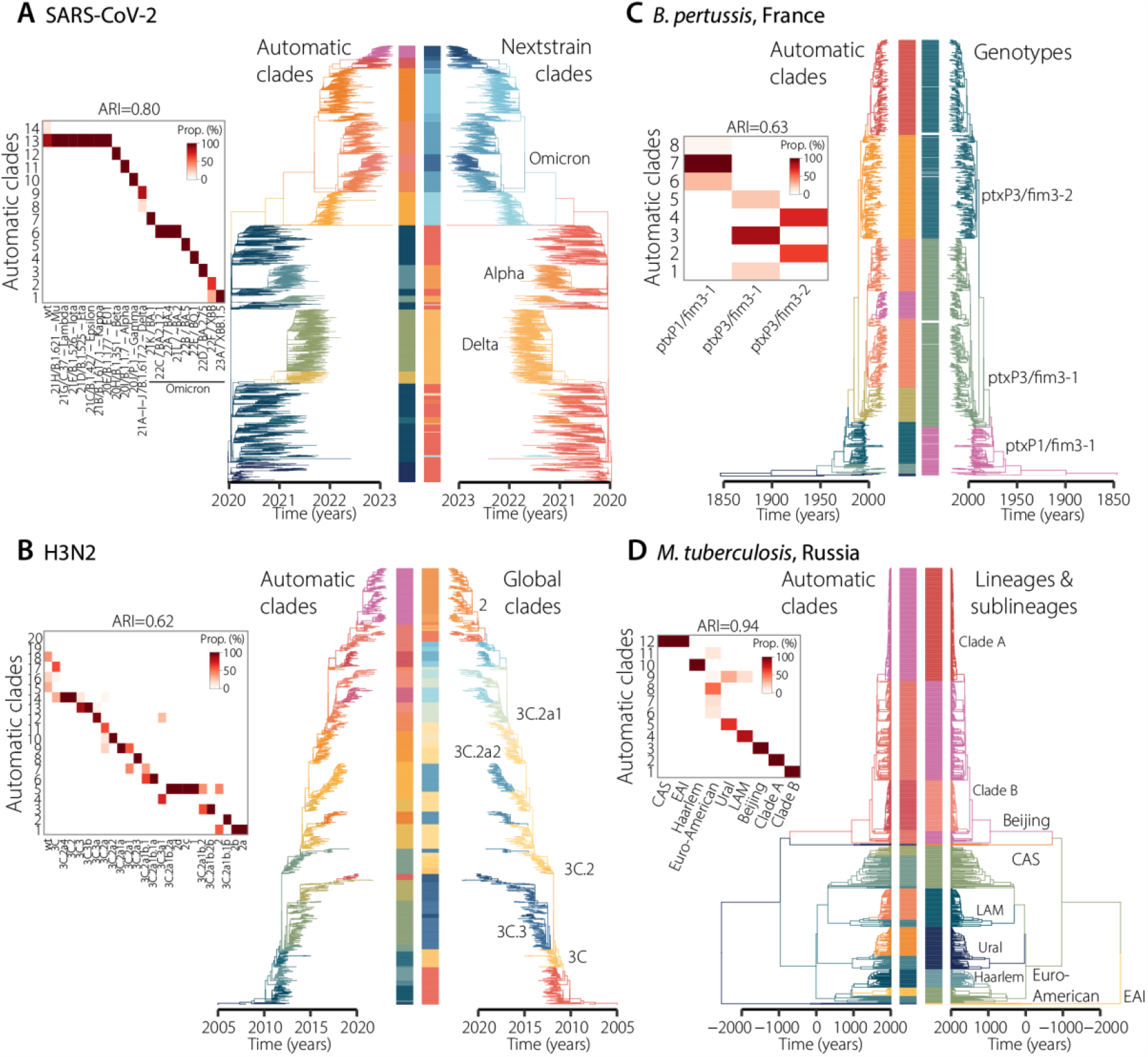
Comparison of the identified lineages to the known population composition. For each pathogen, we present a heatmap comparing the known population structure (x-axis) to the automatic clades found by our framework (y-axis). Darker colors represent more agreement between both classifications. We also compare the timed-resolved phylogenetic trees colored by respective lineage classifications: automatic clades on the left, and previously identified lineages on the right. The colors of the automatic clades are the same as in Figure 1. For *M. tuberculosis*, LAM denotes the Latin American-Mediterranean lineage, EAI denotes the East African Indian lineage and CAS denotes the Central Asian Strain lineage.

### Quantifying the fitness of each detected lineage

We developed a multinomial logistic model that takes into account the birth of lineages to fit the proportion of each lineage through time and quantify their fitness. We assume each lineage has a constant fitness through time, defined as its relative growth rate in the population. By taking into account lineage emergence based on their Most Recent Common Ancestor (MRCA), our model does not estimate proportions for lineages that do not exist yet in the population, as opposed to implementations in other studies, e.g., (*8*). This simple model captured the lineage dynamics of each pathogen (Figure 3A-D and S6-9). We found that the underlying fitness of each emerging lineage was non-null, in line with the lineages called being indeed differently fit (Figure S10). We further computed the inferred real-time fitness of each lineage in the population. Indeed, while our model estimates a constant fitness parameter for each lineage, their actual fitness through time depends on what other lineages are circulating at that time. We found that the SARS-CoV-2 lineage 1, corresponding to Omicron XBB1.5, had the best maximal real-time fitness, followed by lineages 5 and 7, corresponding to Omicron BA.5 and BA.1 (Figure 3E, S10). H3N2 lineages’fitness was more homogeneous across the population, with lineages persisting on average 3.9 years after their emergence (Figure 3F, S10)(*30, 31*). For *B. pertussis*, our results are consistent with those of previous studies(*3*). However, we note that three lineages (labeled 1, 2 and 3) emerged following the implementation of a new acellular vaccine in France in 1998(*32*) (Figure 3G, S10). We found that these three lineages have the highest fitness of all *B. pertussis* strains, pointing towards a potential immune pressure on lineage dynamics from the new vaccine. *M. tuberculosis* lineage fitness was the most stable of the four pathogens explored, reflecting its long-lasting diverse population. The only exception is the comparatively recent emergence of lineages 1 and 2(*20*) (Figure 3H, S10). These lineages are rising sharply in the population, and have a relative fitness per year of 1.0057, 95%CI:[1.0055, 1.0060] and 1.00087, 95%CI:[1.00077, 1.00098], respectively.

**Figure 3:**
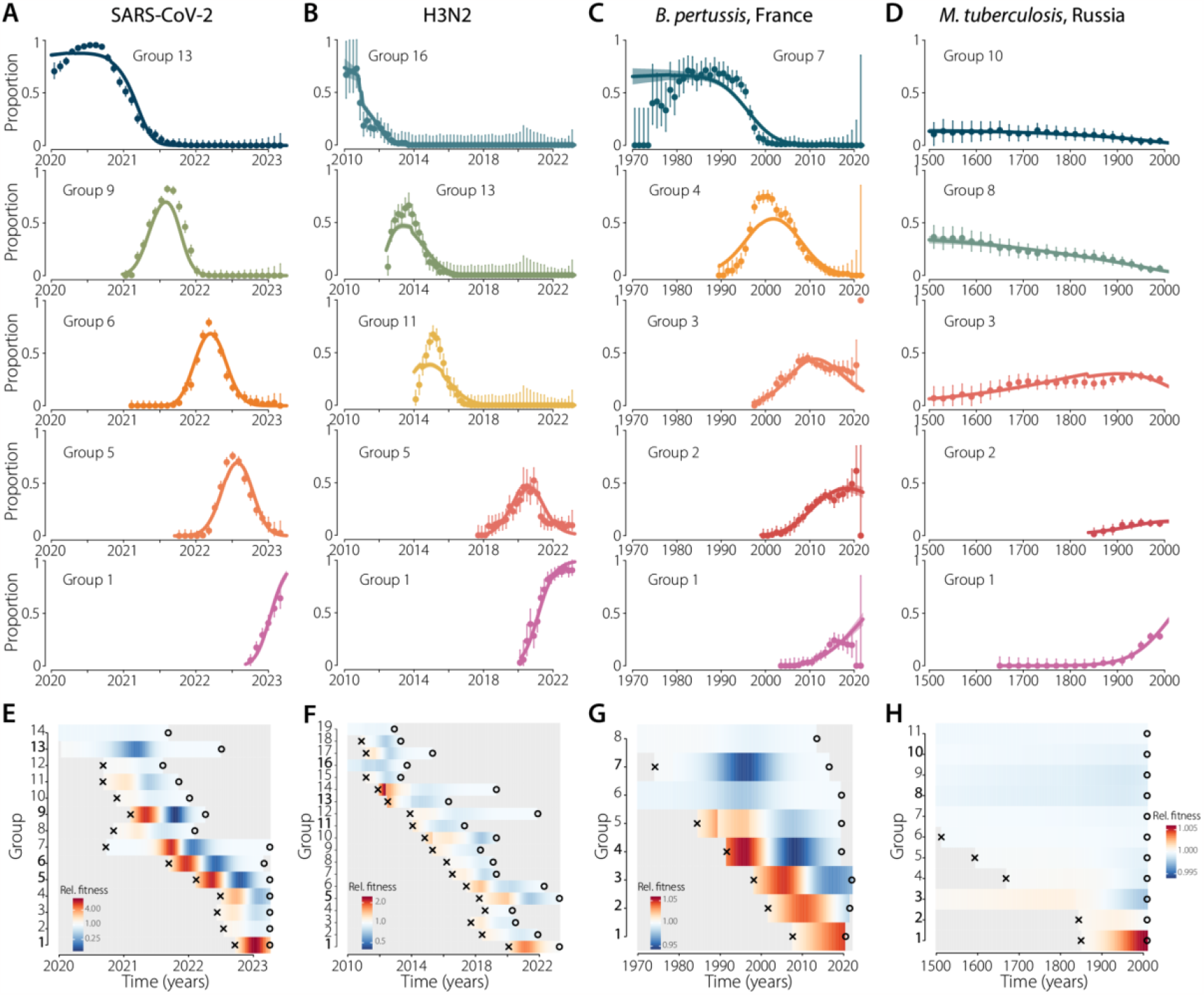
Estimation of the fitness of each lineage. **(A-D)** Model fits per pathogen. For each pathogen, we present the fits for the five most prevalent groups. The fits for all groups are presented in Figure S6-9. Colored dots represent data, bars denote 95% confidence intervals. Colored lines and shaded areas represent the median and 95% credible interval of the posterior. **(E-H)** Relative fitness of each group, over time. Estimates for all groups are presented in Figure S10. Crosses indicate the group’s MRCA. Open circles indicate the last isolate from each group, in our datasets.

### Lineage-defining mutations

We explored whether specific changes in the genomes were linked to lineage fitness by identifying lineage-defining mutations (Figure 4). We defined such mutations as (i) present in at least 80% of the sequences in that lineage and (ii) not present in the ancestral lineage. While we focus on mutations, we note our framework is applicable to other covariates, both for the analysis of genotypes (e.g. indels, or gene gain/loss), or phenotype (e.g. resistance to antimicrobial drugs). For each pathogen, we looked at where those mutations are located in their genomes, and how functionally relevant each of them are. For SARS-CoV-2, we found that the highest density of lineage-defining amino-acid substitutions was located in the Receptor Binding Domain (RBD) of the spike protein, with low densities in ORF1a, ORF1b, and ORF10 (Figure 4A-E-I, S11, S12). Our lineage-defining mutations were consistent with those described in a previous analysis that estimated nucleotide positions linked with shifts in fitness across 6 million SARS-CoV-2 genomes(*8*). We found that our screening recovered the fittest mutations (Figure 4I). We obtained similar results with H3N2, for which most of the lineage-defining amino-acid substitutions are located in the HA1 domain (Figure 4B-F-J, S13). We then investigated specifically if the mutations that we found were located in previously described antigenic sites(*33*). We found that indeed, the antigenic sites had the highest proportion of amino acid substitutions compared to the rest of the gene, and that within those, the Koel sites had the highest proportions of substitutions(*34*) (Figure 4J). Our framework also gave interesting results in *B. pertussis* and *M. tuberculosis*. We recovered the main previously-described pertussis lineage-defining mutations, namely in ptxP and *fim3* (Figure 4C-G-K). Further, we found a selection of other associated mutations that had not been previously described, with two distinct non-synonymous mutations in *sphB1* being of particular interest as they suggest parallel evolution (Figure S14). *sphB1* encodes a protease which is involved in the extracellular release of the pertussis filamentous haemagglutinin, a *B. pertussis* acellular vaccine antigen and key host-interaction factor(*35*). Overall, we found that virulence-associated genes had the highest proportion of lineage-defining mutations (Figure 4K). Lastly, we investigated the mutations associated with the most recent clades of *M. tuberculosis* (clades 1 and 2 from Figure 3H). As reported previously(*20*) we found that antimicrobial resistance-associated genes had the highest proportion of lineage-defining mutations (Figure 4D-H-L, S15).

**Figure 4:**
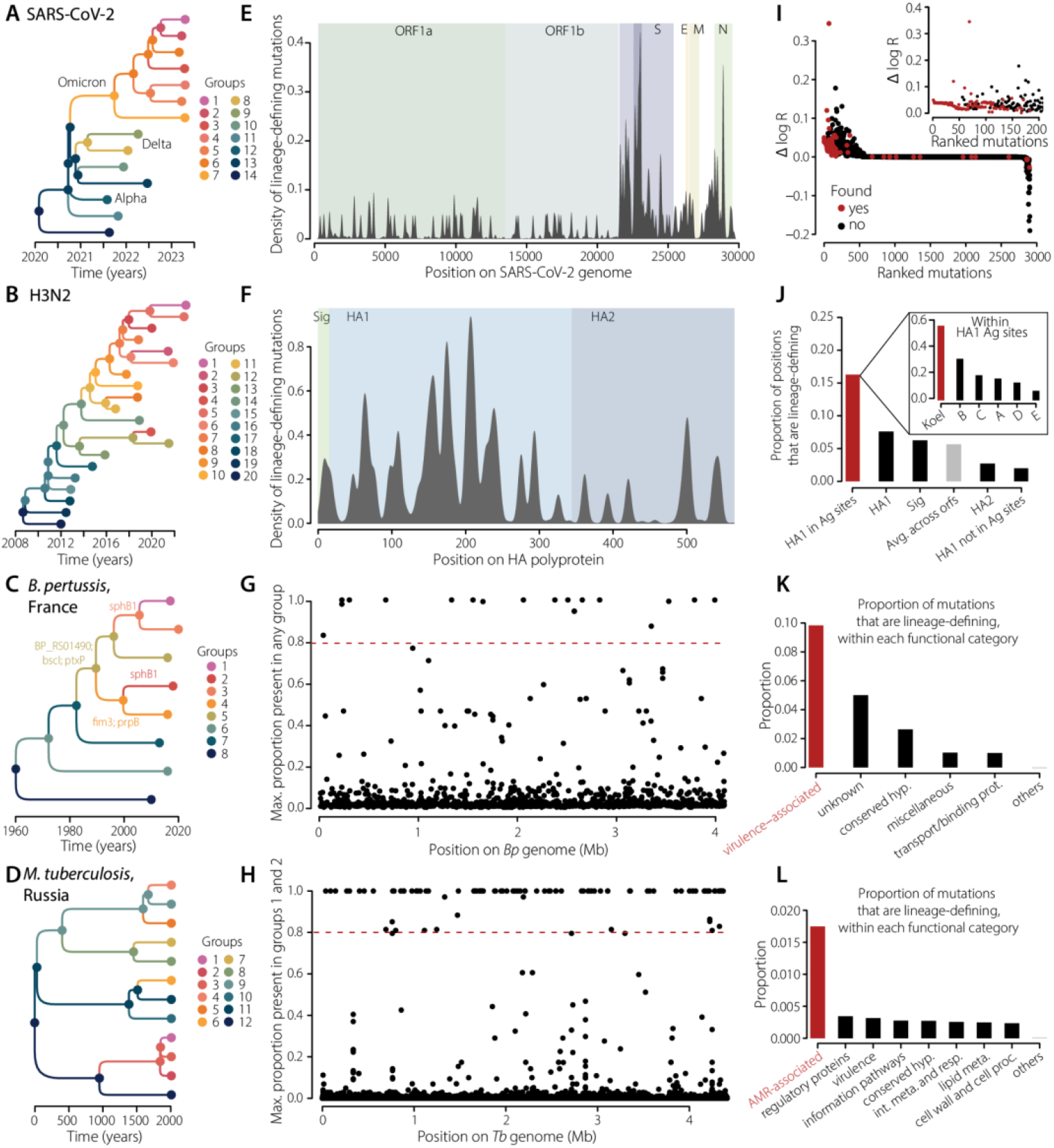
Lineage-defining genetic mutations. For each pathogen, we present a summary of the genetic evolution of the lineages. **(A-D)** For each pathogen, we present the lineage trees representing the genealogical relationship between them. Colors indicate groups. **(E-H)** Lineage-defining mutations along the genome of each pathogen considered. For SARS-CoV-2 (E) and H3N2 (F) viruses we plot the density of lineage-defining mutations along the full genome (SARS-CoV-2) or HA polyprotein (H3N2). Colors indicate the main ORFs. For *B. pertussis* (G) and *M. tuberculosis* (H) we plot for each mutation the maximum proportion of that mutation that is present in any group *(B. pertussis)* or in groups 1 and 2 *(M. tuberculosis)*. The dashed lines represent the 0.8 cutoff. The lists of mutations identified can be found in Data Files S5-8. **(I-L)** Functional relevance of the mutations identified. (I) For SARS-CoV-2, we compare the substitution analyzed by Obermeyer and colleagues(*8*) (black), and the mutations found to be lineage-defining in our study (red). (J) For H3N2, we plot the proportion of positions that are lineage-defining within each HA polyprotein subunit, and antigenic sites(*33, 34*)(insert). (K) For *B. pertussis*, we plot the proportion of mutations that are lineage-defining within each functional category(*36*) (L) Same as K, for *M. tuberculosis(37*). The lists of lineage-defining mutations for each pathogen can be found in DataFiles S5-8.

### Tracking lineages in real-time

Our framework enables us to track population composition changes through time, with a direct link to fitness. As our method relies on the estimation of the pairwise distance distribution for each node in a tree, the number of sequences does not impact the index dynamics, as long as sequences are representative of the diversity (Figure 5A). To demonstrate this robustness to sampling biases in time, we conducted a sensitivity analysis using the SARS-CoV-2 dataset by repeatedly removing a subset of genomes, including in a temporally uneven manner, and re-estimated the circulating lineages each time. We found that our framework was still able to detect virtually all the lineages, even when using heavily biased datasets (Figure 5B, mean ARI of 0.90). Finally, we explored how fast after emergence our framework was able to detect lineages. We truncated our full global SARS-CoV-2 dataset every two weeks and reran the detection algorithm. We found that our model was able to capture each lineage, with a median delay of 2.2 months after emergence, with only 10 sequences required (Figure 5C). Considering that the SARS-CoV-2 dataset used in this study comes from NextStrain and was composed of only 3129 sequences (approximately 0.02% of all sequences available on GISAID at the time of the study), the time to lineage identification could be further shortened.

**Figure 5:**
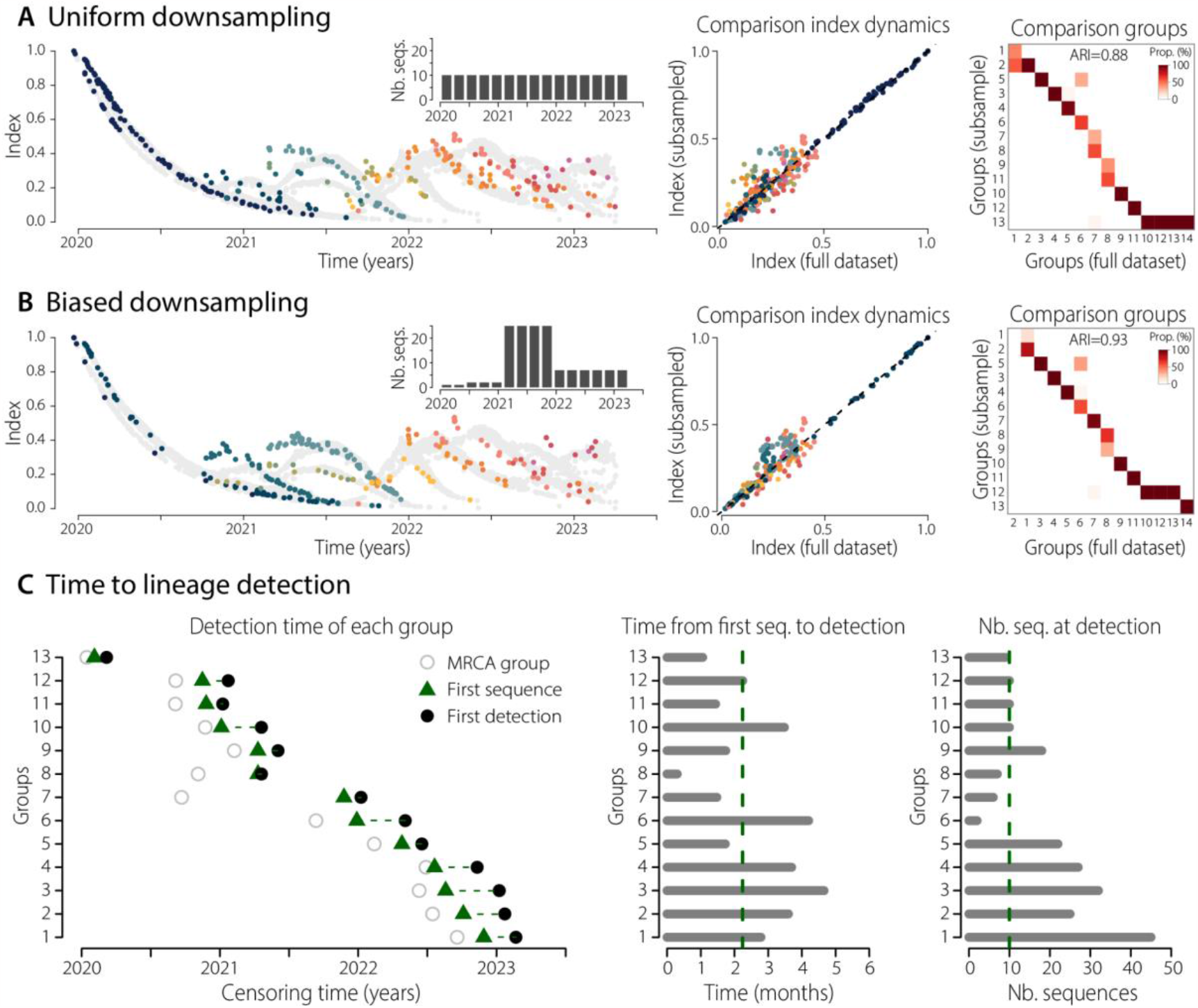
Robustness of the framework to sampling intensities and time to lineage detection. **(A-B)** Robustness to downsampling. We kept only 150 sequences from the global SARS-CoV-2 tree, either sampled uniformly through time (A) or in a temporally uneven manner (B). From left to right: Index dynamics computed on the subsampled trees, colored by detected lineages, with temporal distribution of sequences in inserts; pairwise comparison of the index computed at nodes (internal and terminal) in the trees from the full dataset (x-axis) and subsampled datasets (y-axis); heatmap comparing the automatic clades found by our framework on the full dataset (x-axis) to the automatic clades found on the subsampled datasets (y-axis). Darker colors on the heatmap denote more agreement between both classifications. **(C)** Time to lineage detection. The full global SARS-CoV-2 dataset was censored every two weeks and reran the detection algorithm. From left to right: detection time of each group, with open circles denoting the group’s MRCA in our tree, the green triangles denoting the first sequence of the group in our dataset, and the black dots denoting the first detection of the group by our framework; time from first sequence isolated in our dataset to group detection; number of sequences within each group at the time of detection. The dashed lines denote the median time to detection, or number of sequences at detection, respectively.

## Conclusion

In this study, we presented a novel framework that can agnostically track changes in population composition in phylogenetic trees, even in situations of heavily biased availability of sequences. Across a broad range of pathogens, we have shown we can recover the main known circulating lineages for each pathogen, as well as identify new, previously unknown lineages, with significant changes in fitness. We can quantify the relative fitness of each lineage and identify genetic changes linked to the emergence of new, fitter lineages. This framework can have important implications for public health surveillance. There is increased interest in the systematic sequencing of pathogens detected in healthcare settings. By integrating such sequencing efforts into our framework, public health agencies will be able to identify emergent strains in a timely manner, which can be used to promote targeted interventions. Our framework is also able to make fundamental insights into pathogen ecology. By quantifying the relative fitness advantage of new strains, our framework can help us identify potential drivers of emergence, including the role of population immunity from natural infection or vaccination. Finally, by identifying the specific genomic changes linked to fitness changes, this work provides testable biological hypotheses into genetic variants in each pathogen that are driving the changes in population fitness of that pathogen.

## Supporting information

DataFile S1

DataFile S2

DataFile S3

DataFile S4

DataFile S5

DataFile S6

DataFile S7

DataFile S8

## Data Availability

Code to replicate the main analyses of this paper will be available at https://github.com/noemielefrancq/paper-index-fitness-dynamics-trees. All sequences generated within this study were deposited in ENA, with accession numbers and metadata attached to each sequence in DataFile S3. All sequences used in this study are available online on GenBank and ENA (B. pertussis, M. tuberculosis) or GISAID (H3N2, SARS-CoV-2). Their accession numbers and metadata are listed in DataFiles S1-4.

## Acknowledgements

We thank Caitlin Collins, Megan O’Driscoll, Angkana T. Huang and Trevor Bedford, for useful discussions and feedback. We thank all the contributors to GISAID for sharing their data.

## Author contributions

Conceptualisation: N.L., J.P. and H.S. Method development and modeling analysis: N.L., supported by L.D., J.P. and H.S. Isolate and genomic data collection: N.L., S.B. and V.B. Supervision: J.P. and H.S. Writing – original draft: N.L. Writing – review and editing: N.L., L.D., V.B., S.B., J.P. and H.S. All authors provided input to the manuscript and reviewed the final version.

## Data and materials availability

Code to replicate the main analyses of this paper will be available at https://github.com/noemielefrancq/paper-index-fitness-dynamics-trees. All sequences generated within this study were deposited in ENA, with accession numbers and metadata attached to each sequence in DataFile S3. All sequences used in this study are available online on GenBank and ENA *(B. pertussis, M. tuberculosis)* or GISAID (H3N2, SARS-CoV-2). Their accession numbers and metadata are listed in DataFiles S1-4.

## Supplementary materials

Materials and methods

Figures S1-S22

Data Files S1-S8

## List of supplementary materials

### Materials and methods

#### Figures

Figure S1: Index dynamics colored by known lineages

Figure S2: Lineage detection based on index dynamics for each pathogen

Figure S3: Index dynamics of SARS-CoV-2 across continents

Figure S4: SARS-CoV-2 lineages identified across continents

Figure S5: SARS-CoV-2 lineages identified with treestructure and fastbaps

Figure S6: Fitness model fits for all lineages of SARS-CoV-2

Figure S7: Fitness model fits for all lineages of H3N2

Figure S8: Fitness model fits for all lineages of *B. pertussis*

Figure S9: Fitness model fits for all lineages of *M. tuberculosis*

Figure S10: Fitness estimates for all pathogen lineages

Figure S11: Proportion of mutations that are defining the lineages of SARS-CoV-2 worldwide, by ORFs

Figure S12: Phylogenetic tree and mutations in the spike protein that are defining lineages in the global SARS-CoV-2 dataset

Figure S13: Phylogenetic tree and mutations in the HA1 subunit that are defining lineages in the global H3N2 dataset

Figure S14: Phylogenetic tree and mutations defining lineages in the *B. pertussis* dataset from in France

Figure S15: Phylogenetic tree and mutations defining lineages 1 and 2 in the *M. tuberculosis* dataset from in Samara, Russia

Figure S16: Population history, pairwise distance distribution and index dynamics.

Figure S17: Robustness of the framework to the choice of timescale

Figure S18: Non-explained deviance as a function of the number of groups in the lineage detection algorithm

Figure S19: Proportion of synonymous mutations that are lineage-defining, by gene functional categories, for *B. pertussis* and *M. tuberculosis*

Figure S20: Example of index dynamics on censored global SARS-CoV-2 datasets

Figure S21: Illustration of the index behavior in different population histories

Figure S22: Robustness to sampling schemes, from simulation study

### Data Files

Data File S1: Isolates SARS-CoV-2 Data File S2: Isolates H3N2

Data File S3: Isolates *Bordetella pertussis*

Data File S4: Isolates *Mycobacterium tuberculosis*

Data File S5: List of SARS-CoV-2 lineage-defining mutations Data File S6: List of H3N2 lineage-defining mutations

Data File S7: List of *Bordetella pertussis* lineage-defining mutations

Data File S8: List of *Mycobacterium tuberculosis* lineage-defining mutations

## Materials and methods

### Sequence data

For each pathogen, we compiled a dataset to investigate the changes in the population composition. For SARS-CoV-2 and Influenza H3N2, we extracted the datasets from the publicly available NextStrain timed-resolved phylogenies accessed on 14 April 2023(*38*). These datasets are sub-samples from all publicly available sequences in GISAID, to represent the diversity as much as possible (we used the ‘all-time’dataset for SARS-CoV-2 and the ‘12y’one for H3N2). In all, we have 3129 whole genome SARS-CoV-2 sequences sampled from 26 December 2019 to 3 April 2023, and 1476 Influenza Hemagglutinin (HA) sequences from 1 January 2005 to 3 April 2023 (Data File S1-2). For *B. pertussis*, we used 1248 sequences from 1953 to 2022, collected by the National Reference Center (NRC) for Whooping Cough and Other Bordetella Infections in France (Data File S3). This dataset is composed of 1023 sequences previously published and 225 newly sequenced isolates. The new isolates have been sequenced with the same methods as previously described(*3*). This dataset is representative of the *B. pertussis* diversity in France as the NRC is receiving isolates from 42 sentinelle hospitals throughout France. For *M. tuberculosis*, we used 997 previously published sequences, isolated in 2008-2010 in Samara, Russia(*20*). This dataset is also representative of *M. tuberculosis* sequence diversity at that location as isolates were prospectively collected from individual patients living in the region and representative of the entire population (Data File S4).

### Multi-sequence alignment for each pathogen

We compiled alignments of all sequences being used. For SARS-CoV-2, we used the precomputed multi-sequence alignment provided by GISAID. For H3N2, we aligned all HA sequences using MAFFT(*39*), with default settings. The alignment was then manually checked. For *B. pertussis* and *M. tuberculosis*, we worked from raw reads. Briefly, adapters and barcodes were stripped from the fastq data and the reads were quality filtered and trimmed using a Phred quality threshold score of 30 using Cutadapt(*40*). We checked the quality of each fastq file using FastQC(*41*). Reads were mapped against the complete Tohama I reference genome (Accession number: NC_002929), or the complete H37Rv reference genome (Accession number: NC_000962.3), using BWA-MEM algorithm(*42*), for *B. pertussis* and *M. tuberculosis*, respectively. Extraction of Single Nucleotide Polymorphisms (SNP) was achieved with the GATK HaplotypeCaller, with ERC GVCF settings(*43*). We kept variants that were present in at least 75% of reads, with a Phred quality score higher than 30, a minimum read depth of 5, a minimum mapping quality of 20 and a String Odd Ratio (SOR) of less than 3. We masked all positions that were covered by less than 5 reads. Further, we filtered out regions which are notoriously difficult to map and/or sequence, similarly to previous studies(*3, 44*). Namely, for *B. pertussis* we filtered out repeated regions (IS481, IS1002 and IS1663)(*36*), and phage regions using Phaster(*45*); for *M. tuberculosis*, we filtered out the functional categories “PE/PPE” or “insertion sequences and phages”(*44*). For *B. pertussis*, we also checked for recombination in our alignment using Gubbins(*46*). As a result, we obtained an alignment of 4701 SNPs for *B. pertussis* and 30533 SNPs for *M. tuberculosis*.

### Reconstruction of timed resolved phylogenies

For each pathogen, we obtained timed-resolved phylogenies. For SARS-CoV-2 and H3N2, we used the NextStrain trees, accessed on 14 April 2023(*38*). For *B. pertussis* and *M. tuberculosis*, we reconstructed the timed phylogenies specifically for this study, using the SNP-based alignments. We first built maximum-likelihood trees using IQ-tree(*47*), using a GTR+F+G substitution model. To assess the branch support, we used the ultrafast bootstrap approximation provided in IQ-tree, performing 1000 replicates for each dataset with the bnni option to reduce the risk of overestimating the branch support(*48*).

For *B. pertussis*, the time-tree was reconstructed using BEAST v1.10.4(*49*), under a GTR substitution model(*18*) accounting for the number of constant sites, a relaxed lognormal clock model(*50*) and a skygrid population size model(*51*). Three independent Markov chains were run for 150 000 000 generations each, with parameter values sampled every 10,000 generations. Runs were optimized using the GPU BEAGLE library(*52*). Chains were manually checked for convergence (ESS values > 200) using the Tracer software(*53*). We manually removed a 10% burn-in.

For *M. tuberculosis*, as all sequences were isolated in 2008-2010, we could not infer a clock rate, but instead, we used a previously estimated clock rate(*54*) of 4.6 x 10^−8^ mutations/site/year. We used the software Bactdating(*55*) to perform a bayesian reconstruction of the timed-tree. We used a fixed mean mutation rate, a relaxed clock rate and a constant effective population size. We ran the chain for 10,000,000 iterations and checked for convergence (ESS values > 200).

### *Inde*x definition

We developed an analytical framework that summarizes the changes in population composition in phylogenetic trees at every time point. Our framework builds on a genetic distance-based index, the Timed Haplotype Density (THD)(*16*), that measures the epidemic success of individual sequences in a dataset. This measure is based on the expectation that sequences sampled from an emerging, fitter, lineage will be phylogenetically closer than the rest of the population at that time. We extend this method to track population changes in phylogenetic trees through time.

We define the *Index* of each isolate *i* in its population at time *t* as:

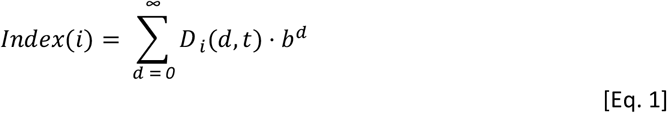

With *D* _*i*_*(d,t)* the distance distribution (in number of mutations or evolutionary time) from the isolate *i* to the rest of the population at that time *t* (Figure 1) and *b*^*d*^, the kernel setting the weight of each distance *d*. *b* is the bandwidth, *b* ∈[*0,1*], which is a parameter to set, linked to the timescale. We compute this index on each node in a tree (internal and terminal).

The weight allows us to track lineage emergence dynamically, focusing on short distances between nodes (containing information about recent population dynamics) rather than long distances (containing information about past evolution). The kernel is governed by the bandwidth *b*, which is a parameter to set. As *b* is dimensionless, it is hard to set. Instead, we use the notion of *timescale* 50 to choose it: the TMRCA such that pairs of isolates with shorter TMRCAs account for 50% of the kernel density(*16*). This timescale is tailored to the specific pathogen studied and its choice will depend on the molecular signal, as well as the transmission rate.

Our definition is virtually the same as the one used by Wirth and colleagues(*16*), with two critical differences: instead of computing the index by summing on each isolate in the population we now sum over the pairwise distance distribution, and we consider the collection time of each sequence to only compute the distance from *i* to the rest of the population that is circulating at that time.

This index is similar to the Local Branching Index (LBI)(*10*), which is defined as total surrounding tree length exponentially discounted with increasing distance from the isolate _*i*_. In our case, rather than considering the tree length, we compute the distance between nodes.

This index definition enables us to write an expectation of the index dynamics over time, as theoretical pairwise distance distributions can be approximated for different populations.

### Linking the Index dynamics to population history

The pairwise distance distribution *D* _*i*_*(d, t)*, or more generally *D (d, t)*, can be seen as the probability, 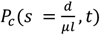, for any pair of sequences sampled at time *t*, to coalesce some time 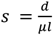 in the past, with *µ* being the rate at which the pathogen accumulates mutations per site and per unit of time, and *l* the length of its genome.

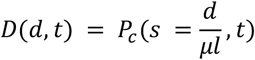

Therefore, at any time point, writing the probability of coalescing in the past enables us to compute the index in the population. We can update equation 1:

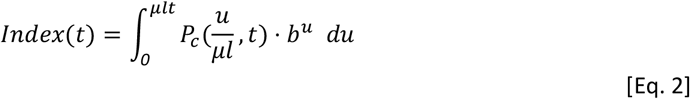

We note that at time *t*, the maximum number of mutations accumulated is equal to *μlt*. For simplicity, we assume a linear accumulation of mutations through time in all the analytical expressions, though one could consider that mutations accumulate randomly given a Poisson distribution with rate *1*/( *µ lt)*.

This probability 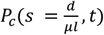 is closely linked to the effective population size. For example, in the simplest case of the structured coalescent process(*17*), if we consider two individuals from a constant population of size *N*_*e*_, we can write their probability of coalescing some time s in the past as:

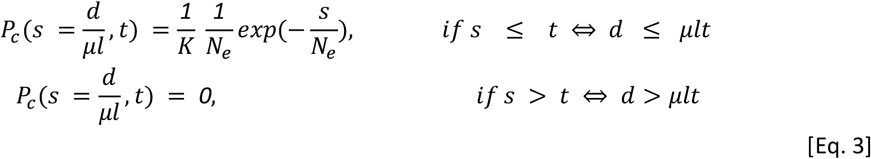

With *K* the normalization constant, so that 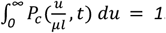.

In Figure S16, we show conceptually how, for different effective population sizes, the probability of coalescing changes, and how it impacts the index dynamics. Formal derivations are presented below in the supplementary text.

### Index computation on timed tree with sequences sampled through time

We use equation 1 to compute the index of each node (internal or terminal) in a timed-phylogenetic tree. To do this, for each node _*i*_, we compute its distance to all the other nodes present in the tree at that time (red crossed on schematic below). All the nodes that fall within the interval of time [*t*_*i*_ −*t* _*wind*_;*t*_*i*_ *+t* _*wind*_] are considered to be circulating at the same time as _*i*_; with *t*_*i*_ being the collection time of the node _*i*_, and *t*_*wind*_ the predefined time window width that is tailored to each pathogen. We also consider extant branches in the computation, as they are evidence of past circulation.

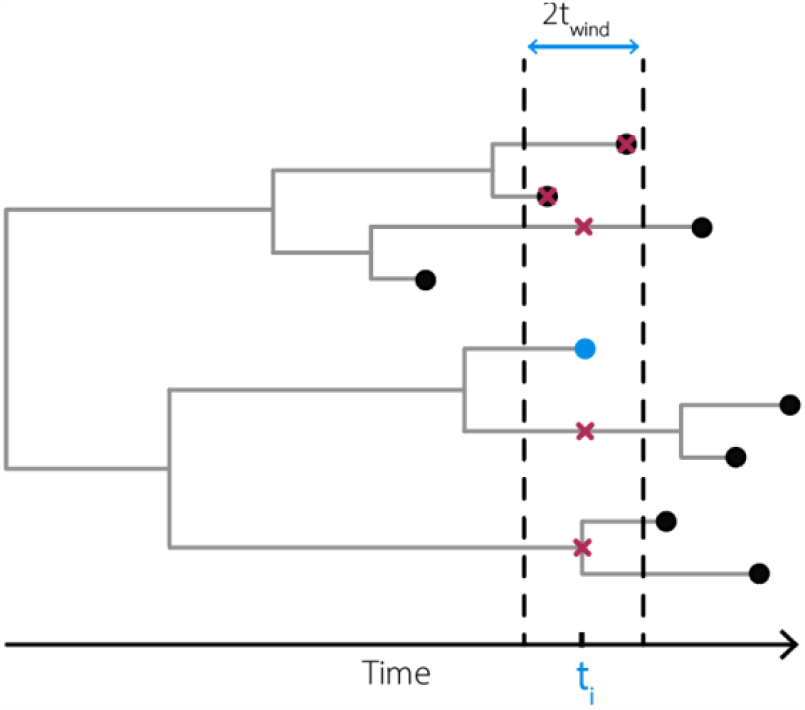

For computation efficiency, similarly to Wirth and colleagues(*16*), we then compute:

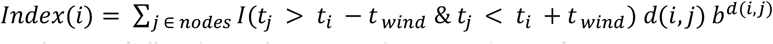

Where *nodes* is the set of all nodes in the tree, and *I* is an indicator function.

This computation is efficient as it only requires i) the precomputation of the indicator function, ii) the precomputation of the distance matrix and iii) a matrix multiplication.

For the pathogens presented in our study we used:

- SARS-CoV-2: a timescale of *0*.*15 years*, and a window of time *t*_*wind*_ = *15 days*
- H3N2: a timescale of *0*.*4 years*, and a window of time *t* _*wind*_ = *0*.*25 year*s
- *B. pertussis*: a timescale of *2 years*, and a window of time *t* _*wind*_ = *1 year*s
- *M. tuberculosis*: a timescale of *30 years*, and a window of time *t*_*wind*_ = *15 years*

We illustrate the impact of the timescale on the index dynamics in Figure S17 on the global SARS-CoV-2 tree.

### Agnostic detection of lineages

We develop a framework that is able to find the set of lineages in the tree that best explains the index dynamics. To do this, we build an algorithm based on generalized additive models (gam) that jointly uses the phylogenetic relationships between nodes in the tree and their index.

In this section, for modelling purposes, we define lineages as monophyletic clades formed by one internal node and all its descendants. Here, these lineages can overlap, meaning that some isolates can be included in multiple lineages. We assume the tree to be binary. For a rooted binary tree with ***n*** terminal nodes, there are ***n***−**2** internal nodes that are not the root, and therefore ***n***−**2** lineages possibilities, which is substantial. To keep the algorithm tractable, we limit the potential list of lineages to those starting with a internal node that has at least ***N***_***off***_ offspring, which is chosen. We note the set of internal nodes to test ***Π***. Further, to increase the accuracy of the detection, we only consider internal nodes that have predefined characteristics:

- For *B. pertussis* and *M. tuberculosis*, as we constructed the bootstrap support of each node (see above), we only consider internal nodes that have a bootstrap support of at least 50% to be the potential start of lineages. This threshold is low, but effectively removes nodes that are not well supported.
- For SARS-CoV-2 and H3N2, instead of bootstrap support, we consider a minimum number of mutations. We only consider internal nodes that have a least 1 mutation on their directly upstream branch.

The log index of each lineage ***l*** is modelled using a cubic spline ***S***_*l*_ (***t, k***) with a pre-defined number of knots ***k***. This allows us to model the log index of each node ***i***, sampled at time ***t***_***i***_, given the lineage that it belongs to:

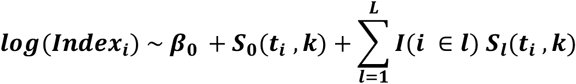

Where β_*0*_ is the intercept, ***L*** is the total number of lineages, *S*_*0*_*(t, k)* and *S* _*l*_*(t, k)* are penalized cubic regression splines with *k* knots(*56*). One ‘null’spline *S*_*0*_*(t, k)* is estimated to model the initial population, together with one spline for each of the ***L*** lineages. If ***L*** = *0*, then no *S*_*l*_ *(t, k)* is estimated. *I ()* is the identity function.

Briefly, the algorithm runs as follows. We start by a null model *M*_*0*_ that fits the index dynamics with one spline *S*_*0*_*(t, k)* (i.e. unstructured population with one single index dynamic, *L* = *0)*. We store the deviance explained *Dev*_*0*_ by the model *M*_*0*_. We then sequentially consider models with increasing complexity *M*_*L*_: we start by first trying models with one lineage, *L* = *1*. We go through the list of internal nodes *Π*that could be the start of a new lineage. When the deviance explained *Dev*_*1*_ by the best model *M*_*1*_ is increased compared to the one of previous null model *Dev*_*0*_, we keep the lineage (effectively the node from *Π)* that explains best the dynamics. We then continue this procedure for increasing *L*. For each number *L*, we go through the list of internal nodes *Π*that could be the start of a new lineage. When the deviance explained *Dev*_*L*_ by the model *M*_*L*_ is increased compared to the one of previous model *Dev*_*L−1*_, we keep the lineage (effectively the node from *Π)* that explains best the dynamics.

The algorithm is implemented in R v4.1.2, using the package mgcv v1.8(*57*) to implement the gam models.

As for any clustering algorithm, choosing the best number of lineages that describe the index dynamics is a challenging question. We took the approach of the elbow plot. We plot the deviances *Dev*_*L*_ explained by each best model *M*_*L*_, as a function of the number of lineages ***L***. This approach enables us to see how well all the models are performing, and to choose the number ***L*** of lineages at which the deviance explained does not increase substantially anymore (Figure S18). From this selected best number of lineages ***L*** _***best***_, we then compute the equivalent set of non-overlapping lineages presented in this paper (Figures 1-5 and S2). We make sure the minimum number of nodes per non-overlapping lineage is at least *N*_*min*_ by merging the small lineages to its closest phylogenetically.

For the pathogens presented in our study we found:

- SARS-CoV-2: 14 lineages, average number of sequences per group of 447, with a set minimum number of *N*_*min*_ = *10*
- H3N2: 20 lineages, average number of sequences per group of 147, with a set minimum number of *N*_*min*_ = *5*
- *B. pertussis*: 8 lineages, average number of nodes per group of 311, with a set minimum number of *N*_*min*_ = *30*
- *M. tuberculosis*: 12 lineages, average number of sequences per group of 181, with a set minimum number of *N*_*min*_ = *30*

To compare the automatic lineages found by our framework to those previously identified, we compute a contingency matrix *C*. Let *U* be the partition of the isolates by our framework, and *V* the partition based on literature. Each element *C*_*i*_,_*j*_ is the number of isolates in both clusters *u*_*i*_ and *v*_*j*_. In Figure 2 we plot this matrix as a heatmap, normalized by column *j*. We computed the Adjusted Rand-Index (ARI) to measure the agreement between partitions, accounting for random clustering(*21*). A value of 1 corresponds to perfect agreement with previously identified lineages, whereas a value of 0 would be expected if clusters were assigned at random.

We illustrate the impact of the timescale on the lineage detection in Figure S16 on the global SARS-CoV-2 tree.

### Quantifying the fitness of each lineage

We developed a multinomial logistic model that takes into account the birth of lineages to fit the proportion of each lineage through time and quantify their fitness.

The proportion *p*_*•,t*_ of sequences at time *t* from each lineage is computed as the number of nodes (internal and terminal) divided by the total number of nodes (internal and terminal) in the population at that time. This proportion *p*_*•,t*_ is modelled by:

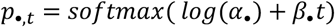

With *α*_*•*_ being the vector of intercept, denoting the initial relative prevalence of each lineage in the population and β_*•*_ the vector of relative growth rates of each lineage. We assume each lineage *i* has a constant relative growth rate β_*i*_ in the population, i.e. each lineage has a constant relative fitness through time. We compute all the relative growth rates with reference to the oldest lineage.

We use a Laplace prior for the growth rate coefficient(*8*):

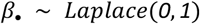

We take into account lineage birth by only allowing *p*_*i*_,_*t*_, the lineage *i* proportion in the population at time *t*, to be non-negative after the lineage’s Most Recent Common Ancestor (MRCA). Formally, this is done by parameterizing *α*_*•*_ as follows. We divide the lineages into two types, either ‘ancestral’, or ‘non-ancestral’:

- An ‘ancestral’lineage is a lineage that is present at the beginning of the time series considered. The total number of ancestral lineages in noted *G*. For those lineages, we sample directly their starting proportions with prior:

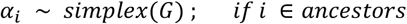
- A ‘non-ancestral’lineage is a lineage that appears after some time - for example the Omicron variant. For those lineages, we assume that their starting frequency, at the time of emergence, is a function of the proportion of their parents in the population at that time. Thus, we write:

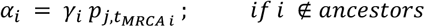

Where *j* is the parent lineage of lineage 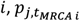 is the proportion of the parent lineage *j* at the time emergence *t*_*MRCAi*_ of the offspring lineage *i*, and *γ*_*i*_ is the share of the parent lineage that is becoming the new lineage. We sample *γ*_*i*_ with a strong prior as we expect that the starting proportion of new lineages should be small:

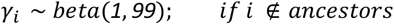

Finally, we update the parent *j* proportion as follows:

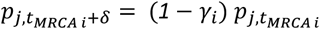

While this parameterization is more complex than the previous efforts using a similar model(*8*), it enables us to take into account that lineages appear through time, which make the model more biologically relevant (e.g., by not estimating the proportion of Omicron in the population in 2020). We chose to parametrize the starting proportions of the new lineages as a function of their parent’s proportions so that i) the model is biologically sound, i.e. the starting proportion of a new lineage cannot be greater than the one of its parent, and ii) the starting proportions are constrained by the proportion of their parents, which makes is statistically easier to fit.

We use a multinomial likelihood to fit the count of sequences per lineage through time *y*_*•,t*_ :

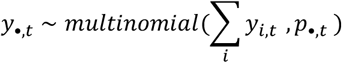

We further computed the inferred real-time growth rate (i.e. fitness) *r*_*i*_ *(t)* of each lineage *i* in the population (Figure 3E-H), to control for the varying presence of all circulating lineages through time. Indeed, while our model estimates a constant fitness parameter for each lineage, their actual fitness through time depends on what other lineages are circulating at that time.

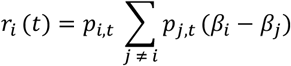

These results are more useful compared to the usual presentation of the parameters, which by default display the relative fitness compared to the ancestral lineage, in this case 19A (the lineage that includes the first SARS-CoV-2 sequences isolated in Wuhan, China).

The model was implemented in Stan, using the *cmdstanr* package(*58*). We ran this model on 3 independent chains with 1,000 iterations and 50% burn-in for each pathogen. We used 2.5 and 97.5 quantiles from the resulting posterior distributions for 95% credible intervals of the parameters.

We fit the counts per lineage in windows of 1 month for SARS-CoV-2, 0.2 year for H3N2, 1 year for *B. pertussis* and 20 years for *M. tuberculosis*, with *t* counted in years for all pathogens.

### Defining mutations of each lineage

We explored whether specific changes in the genomes were linked to lineage fitness by identifying lineage-defining mutations. We defined such mutations as:

- Mutations that are present in more than 80% of the nodes in that lineage
- While those mutations are not present in the set of defining mutations of the ancestral lineage.

For all pathogen, we reconstructed the mutations at each node in the trees using the ancestral state reconstruction implemented in the library ape. To maximize the correct assignment for nodes, we only consider nodes for which the state’s probability was >0.9. Mutations were then classified as synonymous, non-synonymous, or extragenic. For *M. tuberculosis* and *B. pertussis* we also classified each mutation by functional category(*36, 37*).

We computed the density of lineage-defining mutations along the SARS-CoV-2 full genome and H3N2 HA polyprotein with a kernel density estimate (Figure 4E-F). We used a gaussian kernel with a bandwidth of 50 base pairs (bp) for SARS-CoV-2, and a bandwidth of 2.5 amino acid (AA) for H3N2. For *B. pertussis* and *M. tuberculosis* we plot for each mutation the maximum proportion of that mutation that is present in the set of groups considered.

To assess the function relevance of the mutations identified for each pathogen (DataFiles S5-8), we compared them to the literature.

For SARS-CoV-2, we matched the amino acid substitution we found to the ones that Obermeyer and colleagues analyzed(*8*). The authors analyzed 6.4 million genomes up to January 20, 2022 and estimated the fitness effect of 2904 substitutions. Although our global dataset is from an extended period of time (up to 3 April 2023), 84% (N=156) of the lineage-defining mutations were analyzed by Obermeyer and colleagues.

For H3N2, we computed the proportion of positions that are lineage-defining within each HA polyprotein subunit, and antigenic sites(*33, 34*). A position is lineage-defining if it has at least one AA substitution that is lineage-defining. The proportion is computed as follows:

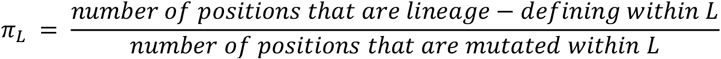

Where L is the set of positions to be analyzed (subunits or antigenic sites).

For the bacteria *B. pertussis* and *M. tuberculosis* we employ a similar metric, by grouping mutations by gene functional categories. We compute:

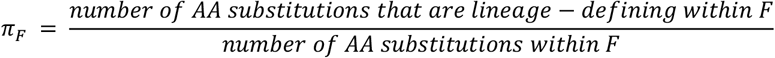

Where *F* is the gene functional category considered(*36, 37*). As a sensitivity analysis, we also replicated this computation on synonymous nucleotide changes, as we expect these mutations to be neutral, and therefore not linked to any particular functional category (Figure S19). We found that, indeed, there was no particular functional category that had significantly more lineage-defining synonymous mutations than others, for both bacteria.

To further check our findings visually, we plotted the lineage-defining mutations for each pathogen next to their phylogenetic trees (Figures S12-15). To make sure the figures were interpretable, we plotted only the mutations in the spike protein for SARS-CoV-2 (Figure S12), the HA1 subunit for H3N2 (Figure S13), and the mutations defining lineages 1 and 2 for *M. tuberculosis* (Figure S14). For *B. pertussis*, we plotted all mutations (AA substitutions and promoter mutations) (Figure S15).

### Robustness to sampling strategies

To demonstrate the robustness to sampling biases in time, we conducted a sensitivity analysis using the global SARS-CoV-2 dataset. We selected two random sets of 150 sequences from the 3129 sequences in our full dataset. We selected them either uniformly through time, or in a temporally uneven manner. To do so, we divided the sequences in 15 time-windows of equal length (79 days). For the uniform sampling, we included 10 sequences per time bin, random selected. For the biased sampling, we included the following number of sequences per bin (see insert on Figure 5B):

- windows 1 and 2: 1 sequence per bin;
- windows 3 to 5: 2 sequences per bin;
- windows 6 to 9: 25 sequence per bin;
- windows 10 to 15: 7 sequences per bin.

After selecting the sequences, we pruned from the tree the ones that were not selected. We then performed the same analysis as described above. We also compared the groups found.

### Analysis of time to detection

We explored how fast after emergence our framework was able to detect lineages. To do this we truncated our full global SARS-CoV-2 dataset every two weeks. Overall, we obtained 81 datasets. Two examples of the index dynamics on censored data on 2021.26 and 2022.50 are presented in Figure S20. We then re-ran the detection algorithm on each dataset. To obtain the best set of lineages automatically for each dataset, we chose the set at which the log deviance explained did not increase by more than 0.01%.

### Simulation study to assess validity of our approach

To demonstrate the validity of our framework, we simulate trees for different population structures. We use the *sim2*.*bd*.*origin* function from the TreeSim package(*59*). It simulates trees based on a birth-death model, with set rates of speciation (birth, *λ)* and extinction (death, *μ)*. A constant effective population size can be simulated by *λ* = *μ*. An exponentially growing effective population can be simulated by *λ >μ*. To simulate a tree with an emerging lineage, we first simulate separately two trees, one with constant effective population size, and one with an exponentially growing effective population size. Then, we randomly select one tip from the first tree and use this tip as the root of the second tree.

In Figure S21, we present those simulations, for three types of effective population sizes: constant, growing, and structured with an emerging lineage. We compare the simulation obtained with the formal expected dynamics (see derivations below). Overall, the simulations verify the validity of our approach. Parameters used: time window: 2 years, timescale: 1 year, mutation rate: 4 mutations per year.

We also reproduced sampling bias to check that our formal expected dynamics are correct even in that case. We sampled the sequences generated either taking 10% of the sequences from year 2-8 or only sequences from years 4-6 and 8-10 (and not years 1-3 or 6-8), mimicking common surveillance system biases. In Figure S22, we present those simulations, with 50 replicates each time. Overall, the simulations verify the validity of our approach. Parameters used: time window: 2 years, timescale: 1 year, mutation rate: 4 mutations per year.

### Expected behavior of the index in a *constant effective population size*

In the simplest case of the structured coalescent process(*17*), if we consider two individuals from a population of constant size *N*_*e*_, we can write their probability of coalescing some time sin the past as (Figure S12C):

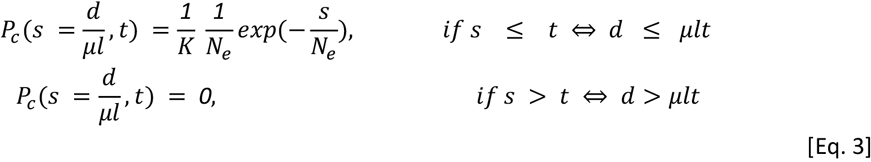

With *K* the normalization constant, so that 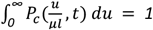.

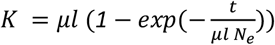

We can plug Equation 3 in the index definition from Equation 2, making sure we take 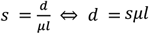. After simplification it follows that:

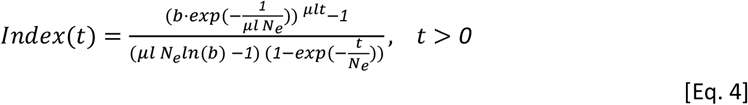

Which is the behavior of the index as a function of time, in a constant population size.

### Expected behavior of the index in a *varying population size*

Following the work of Griffiths and Tavaré (*18*) on the coalescent process in varying population sizes, we can further derive the index in more complex population dynamics. We set the effective population size of our lineage to ***N***_***e***_(**t**), which can vary through time. We can define the population-size intensity function ***Λ*** by (*18)(19*):

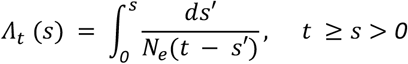

We assume that *Λ(∞)* = *∞*, so that each pair of individuals may be traced back to a common ancestor with probability one (*18*). The density *λ* of *Λ* is given by (*18*):

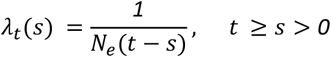

It follows that *P*_*c*_*(s, t)*, i.e. the probability of waiting stime to have the first coalescent event is:

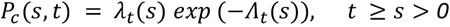

We can find back Equation 3, by taking 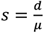 and plugging in a constant population size *N*_e_*(t)* = *N*_e_:

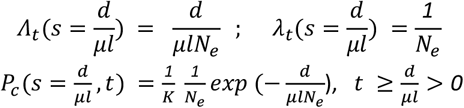

With K the normalization constant. Next, we consider the case of exponentially varying population size.

### Expected behavior of the index in an *exponentially growing effective population size*

We set: *N*_e_*(t)* = *N*_*0*_ ·e^*rt*^, with *N*_*0*_ the initial population size and *r* the rate at which the population is growing (Figure S12F). We assume *r >0*. We can then define the new *λ*_*t*_*(s)* and *Λ*_*t*_*(s)*:

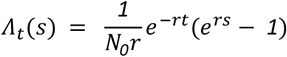

And:

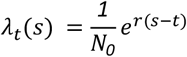

So that:

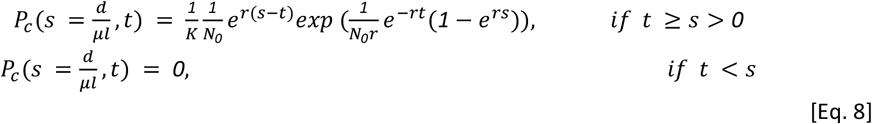

With *K* the normalization constant so that 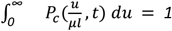

Therefore, we can plug Equation 8 in the index definition from Equation 2, which leads to:

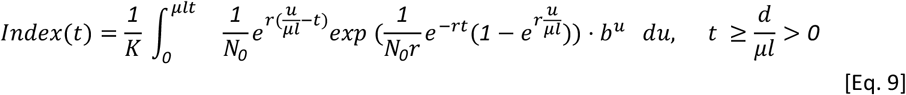

This sum does not have a closed-form expression. However, it can be numerically approximated (Figure S12H).

### Expected index behavior for newly emerging lineage

We can note that in the case of a varying population size (e.g. exponentially varying), the index is dependent on *r*, the rate at which the population size is varying.

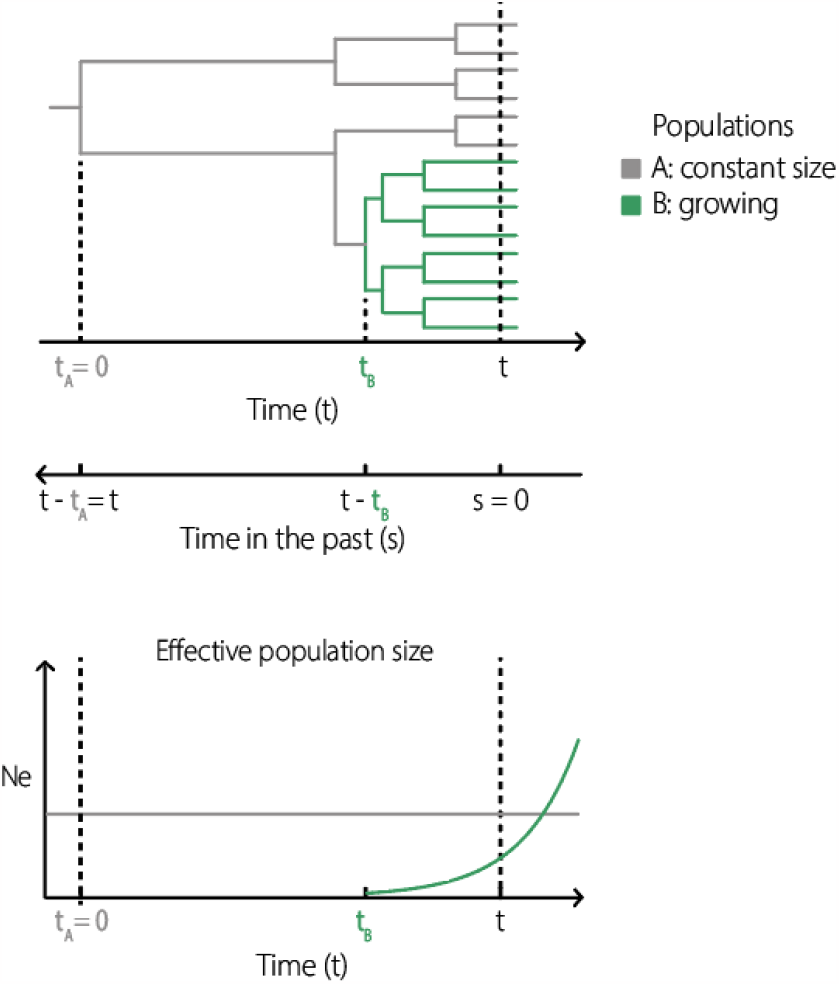

We can derive the index in structured populations that are more complex. For example, we consider here the case of a new lineage expanding in a population (schematic below). Let *Pop*_*A*_ be the ancestral population (schematic below, in gray), with constant effective population size *N*_*A*_, and *Pop*_*B*_, an offspring from *Pop*_*A*_(schematic below, in green), which appeared at time *t*_*B*_. At time *t*_*B*_, the effective population size *N*_*B*_ *(t)* of *Pop*_*B*_ is 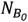. We assume that the *Pop*_*B*_ is growing exponentially 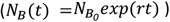 through time with rate *r >1*.

We now write the index of each population. We assume that the appearance of population B has a negligible impact on the index of the individuals sampled from population A. The effective size of population A is constant through time, therefore we can use Equation 4:

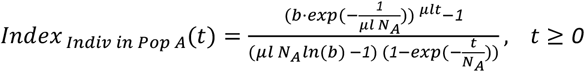

Population B is growing exponentially, within population A, therefore writing the index of individuals sampled from this population is more complex. Let’s consider an individual sampled from population B. Its probability to coalesce with the rest of the population can be separated in two cases:

- It coalesces with an individual from population B, with probability *P*_*c*_,_*B →B*_ *(s, t)*
- Or it coalesces with an individual from population A, with probability *P*_*c*_,_*B →A*_*(s, t)*

The total population through time is: *N*_*tot*_*(t)* = *N*_*A*_ *+N*_*B*_ *(t)*.

Therefore, the probability of an individual sampled from population B to coalesce with another individual in the population is:

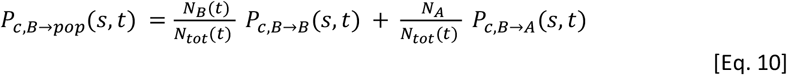

We can note that *P*_*c*_,^*B B*^ *(s, t)* exists only for *t >t*_*B*_ (otherwise population B does not exist yet) and *t* −*t*_*B*_ ≥s ≥*0*, and *P*_*c*_,_*BA*_*(s, t)* exists only for s≥*t* −*t*_*B*_.

First, let’s write *P*_*c*_,_*B →B*_ *(s, t)*. As population B is growing exponentially, we can re-use Equation 9:

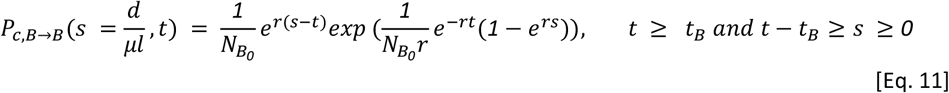

Second, let’s write *P*_*c, B →A*_*(s, t)*. We note that this probability only exists for s≥*t* −*t*_*B*_, and the size of population A is constant. So we can rescale this probability:

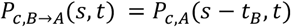

We can note that we already wrote this probability earlier in equation 3, so it follows that:

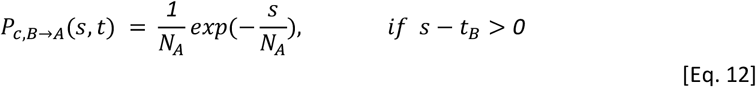

We can now plug Equations 11 and 12 into Equation 10, to obtain the index of individuals sampled from population B:

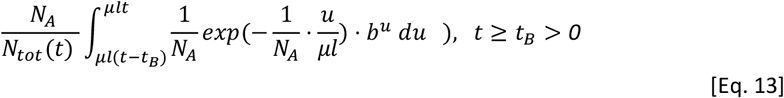

With *K* the normalization constant so that 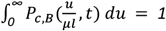

Similarly to Equation 9, this Equation does not have a closed-form expression. However, it can be numerically approximated. Further, we can note that considering only two different populations already makes the index mathematically hard to track, at least without simplifying assumptions.

## Supplementary figures

**Figure S1:**
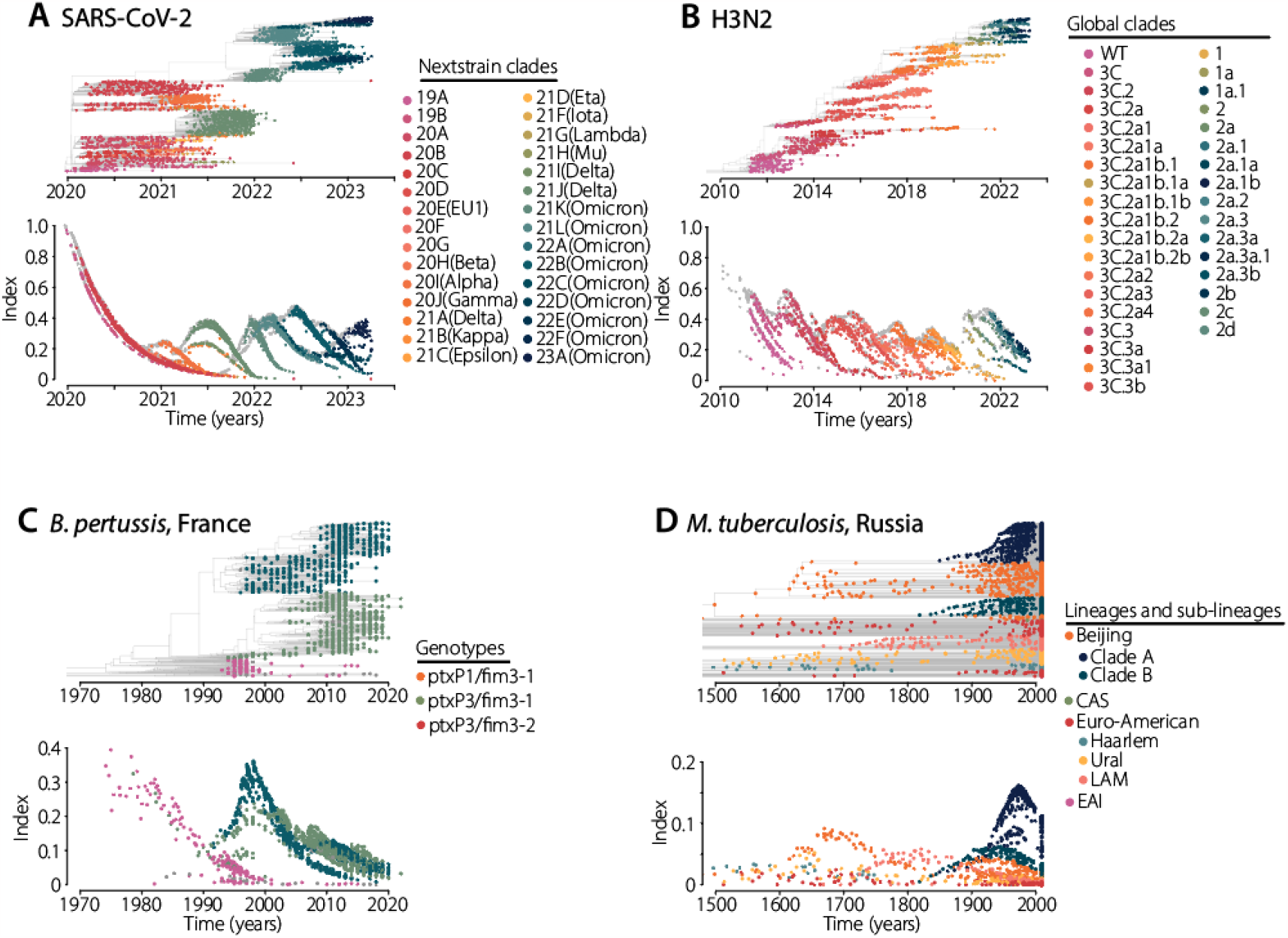
Index dynamics colored by known lineages. Similarly Figure 1B-E, for each pathogen, we present the index dynamics computed at each node (terminal or internal). Here colors represent the different known clades, genotypes, or lineages (see legend on the side). For *M. tuberculosis*, LAM denotes the Latin American-Mediterranean lineage, EAI denotes the East African Indian lineage and CAS denotes the Central Asian Strain lineage.

**Figure S2:**
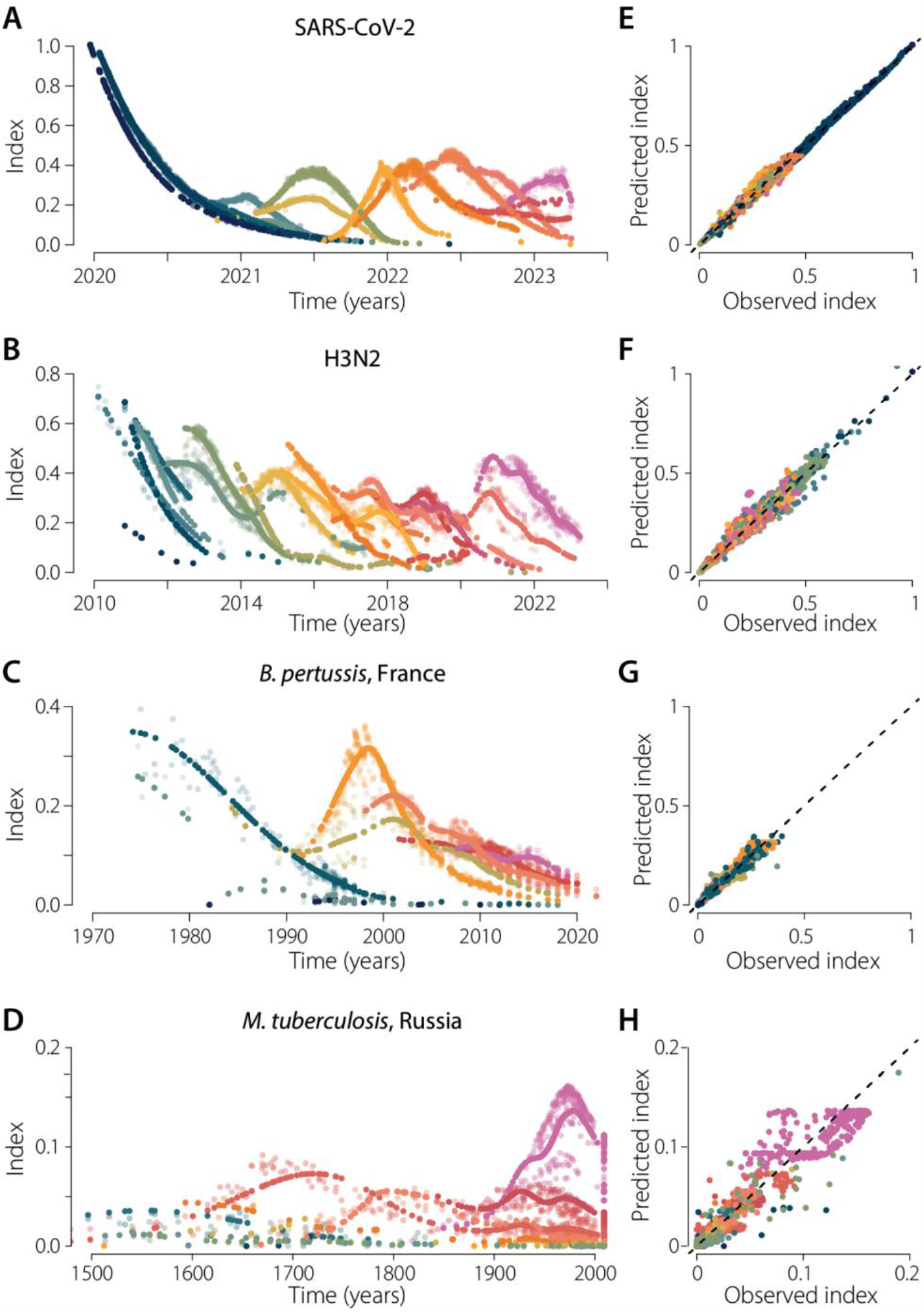
Lineage detection based on index dynamics for each pathogen. **(A-D)** For each pathogen we present model fits of the index dynamics using the best set of lineages. Solid dots represent the model prediction. Shaded dots represent the data. **(E-F)** Predicted versus observed index. The dashed lines denote identity lines. For each pathogen, colors represent the different lineages identified by their different index dynamics (same colors as in Figures 1-4).

**Figure S3:**
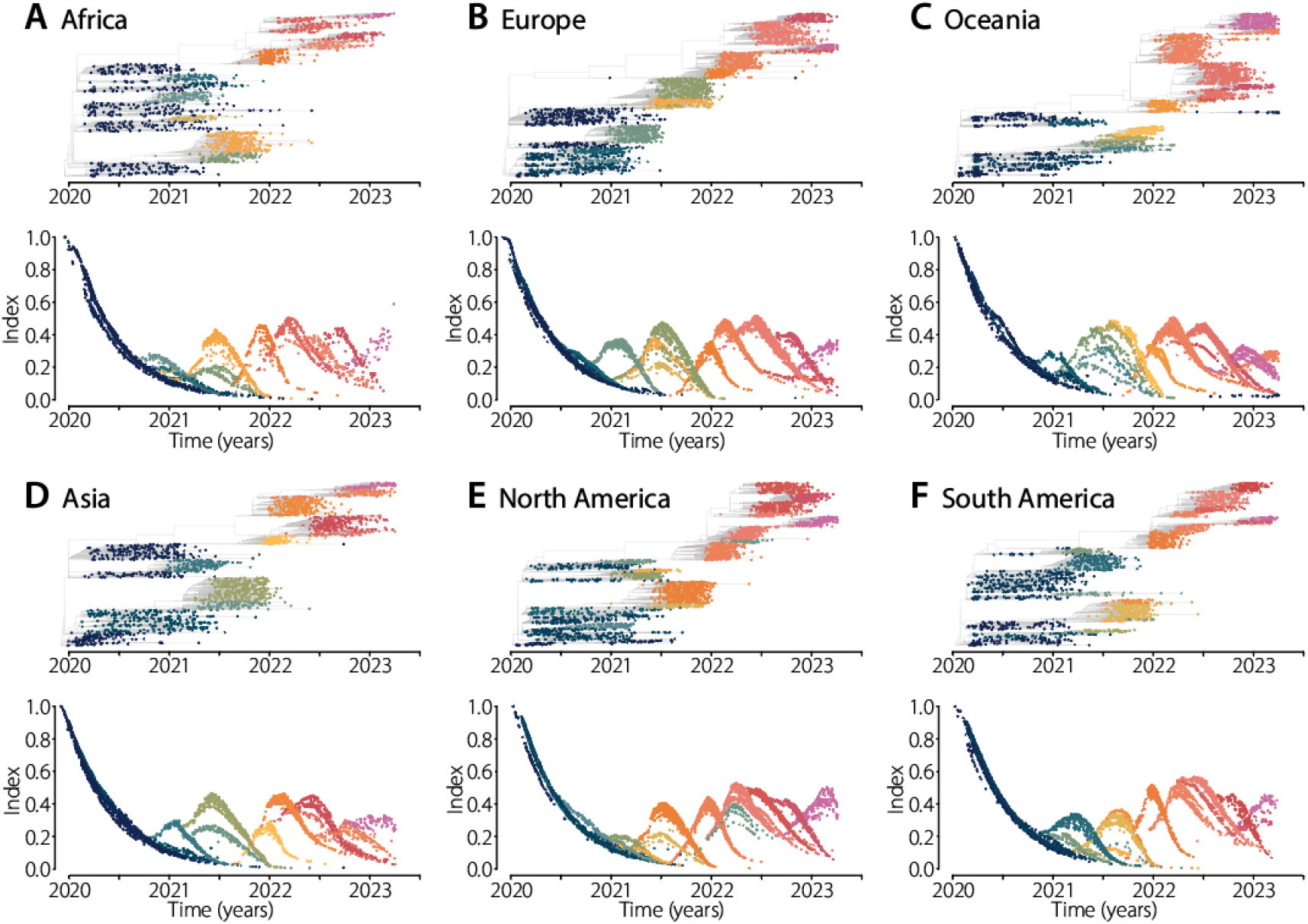
Index dynamics of SARS-CoV-2 across continents. For each continent, we present the index dynamics computed at each node (terminal or internal). Colors represent the different lineages identified by their different index dynamics. Timed-resolved phylogenies for each continent were obtained from NextStrain, accessed on 14 April 2023(*38*).

**Figure S4:**
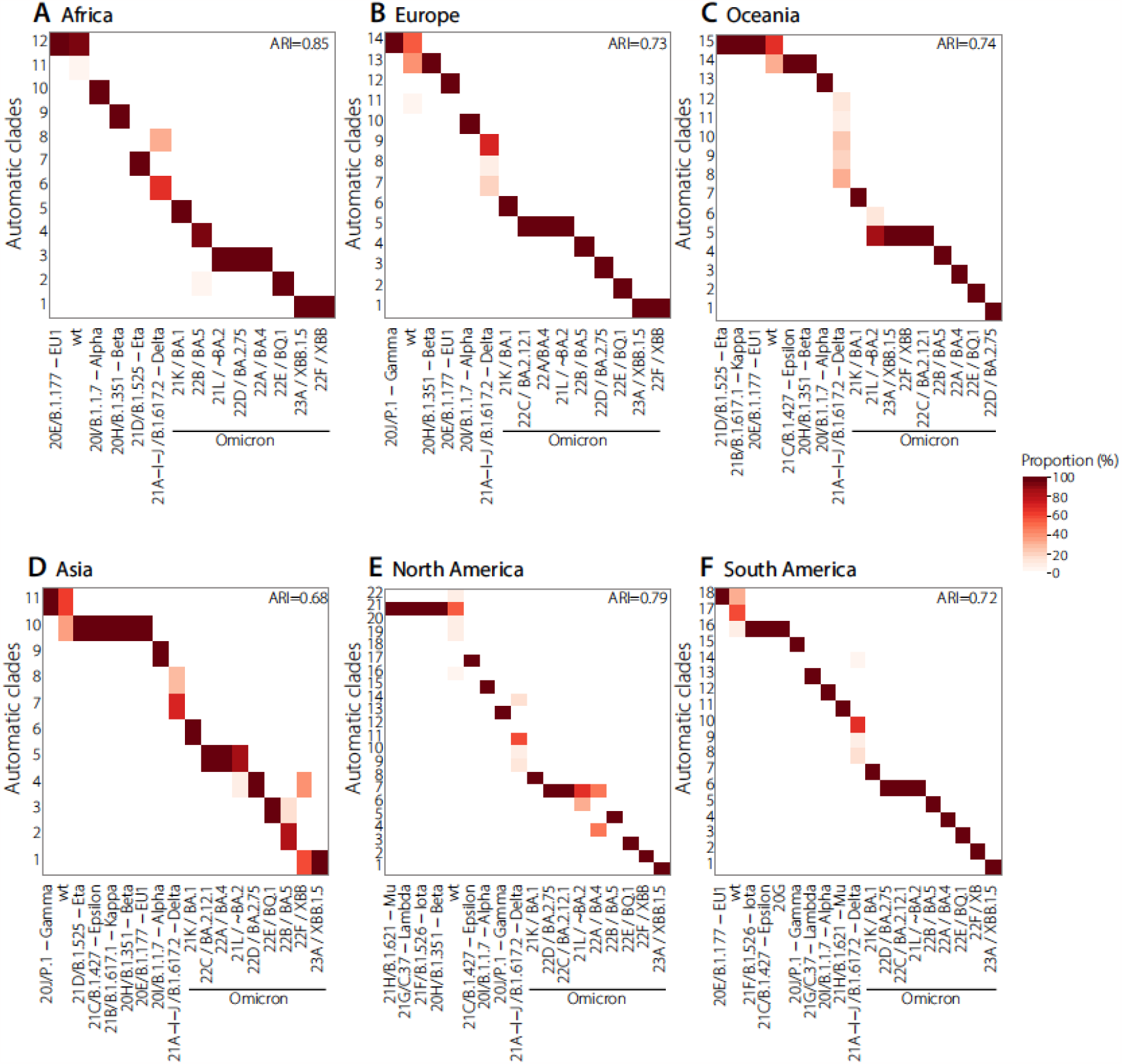
SARS-CoV-2 lineages identified across continents. For each continent, we present a heatmap comparing the known clades identified by NextStrain (x-axis) to the automatic clades found by our framework (y-axis). Darker colors represent more agreement between both classifications.

**Figure S5:**
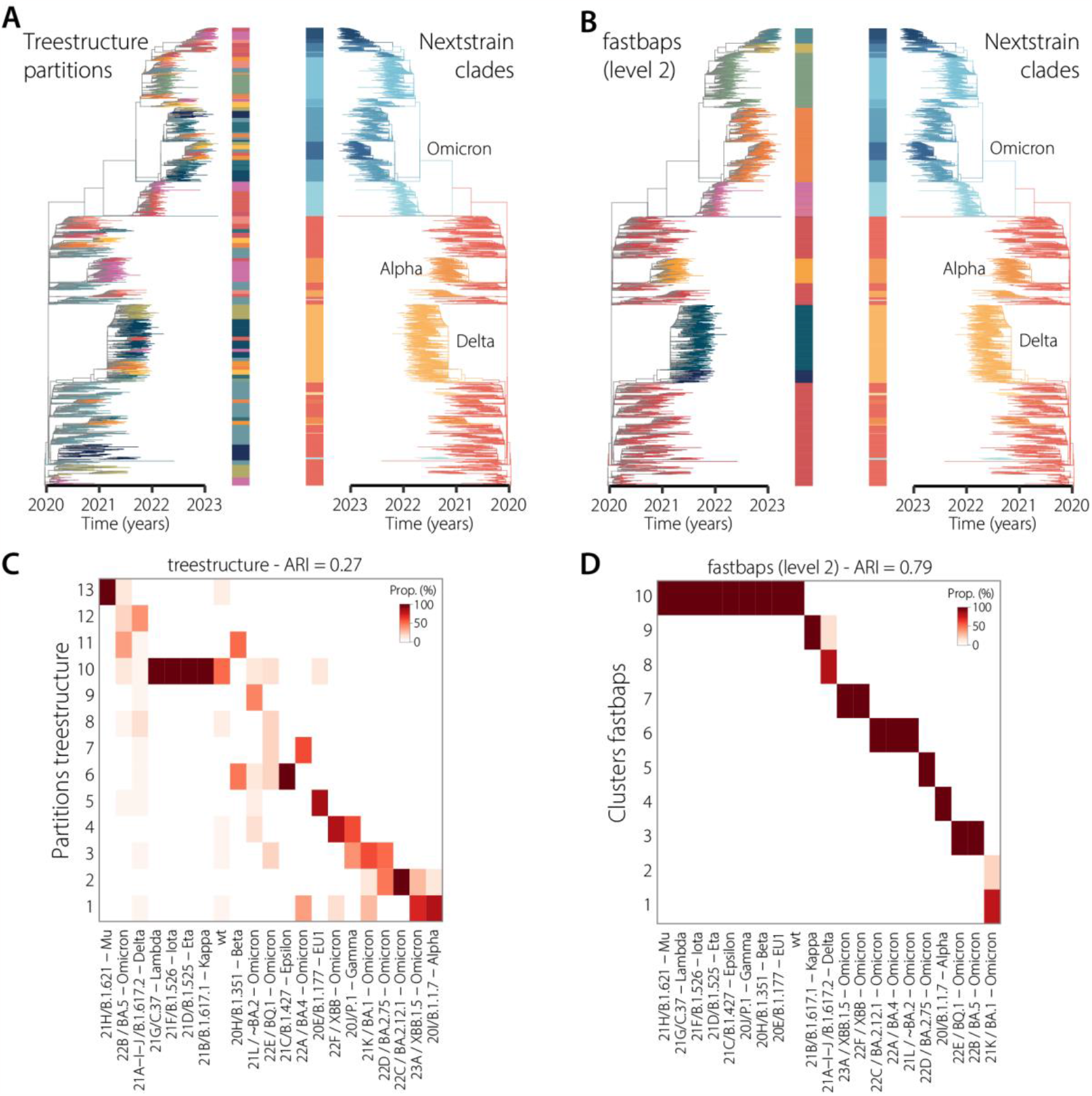
SARS-CoV-2 lineages identified with treestructure and fastbaps. **(A-B)** Global SARS-CoV-2 trees colored by the lineages identified with treestructure (A), or fastbaps (B). **(C-D)** We compare the lineages identified with either algorithm (y-axis) to the NextStrain clades (x-axis). Darker colors represent more agreement between both classifications.

**Figure S6:**
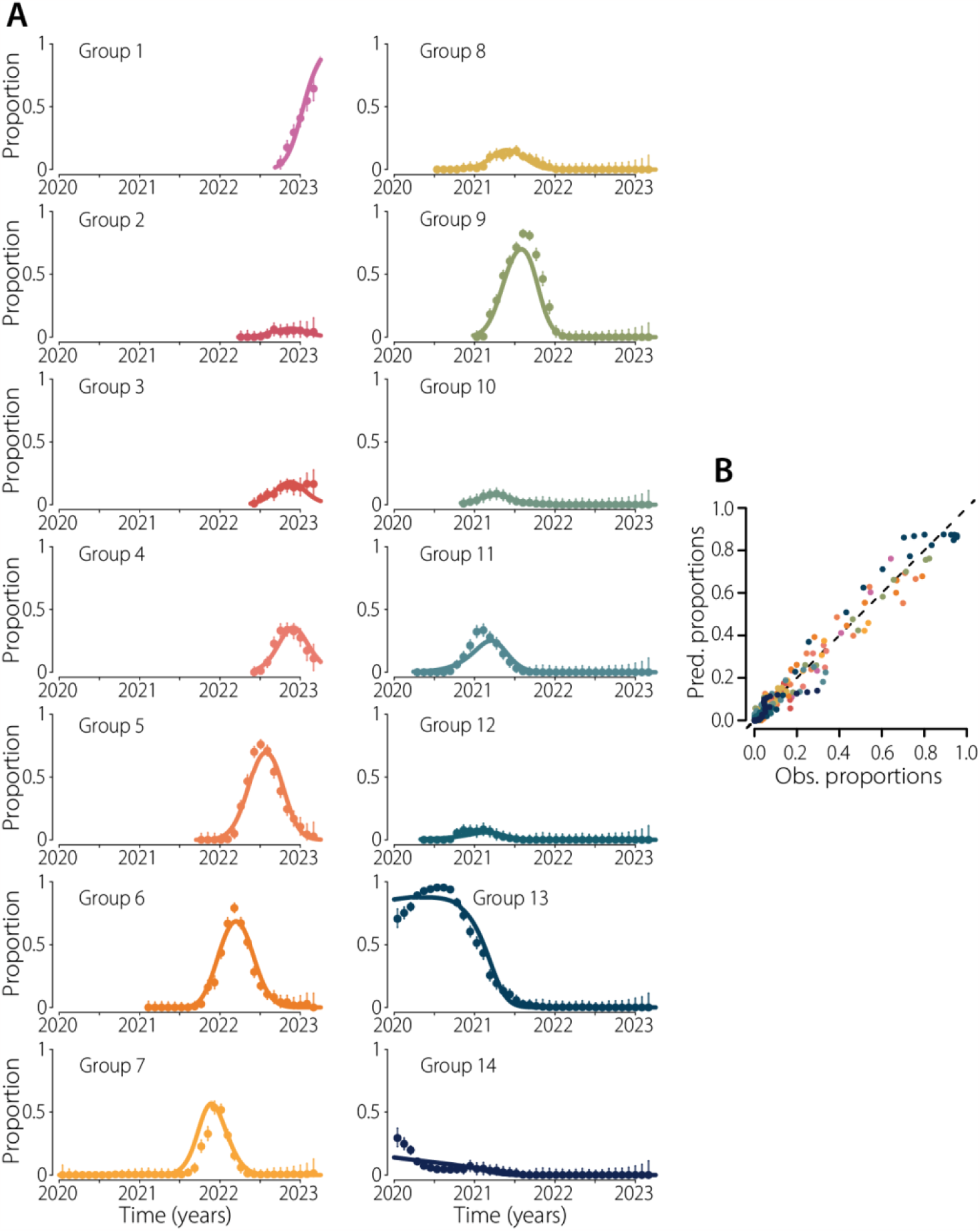
Fitness model fits for all lineages of SARS-CoV-2. **(A)** Fits of the proportion of all the SARS-CoV-2 lineages. Colored dots represent data, bars denote 95% confidence intervals. Colored lines and shaded areas represent the median and 95% credible interval of the posterior. **(B)** Predicted versus observed proportions.

**Figure S7:**
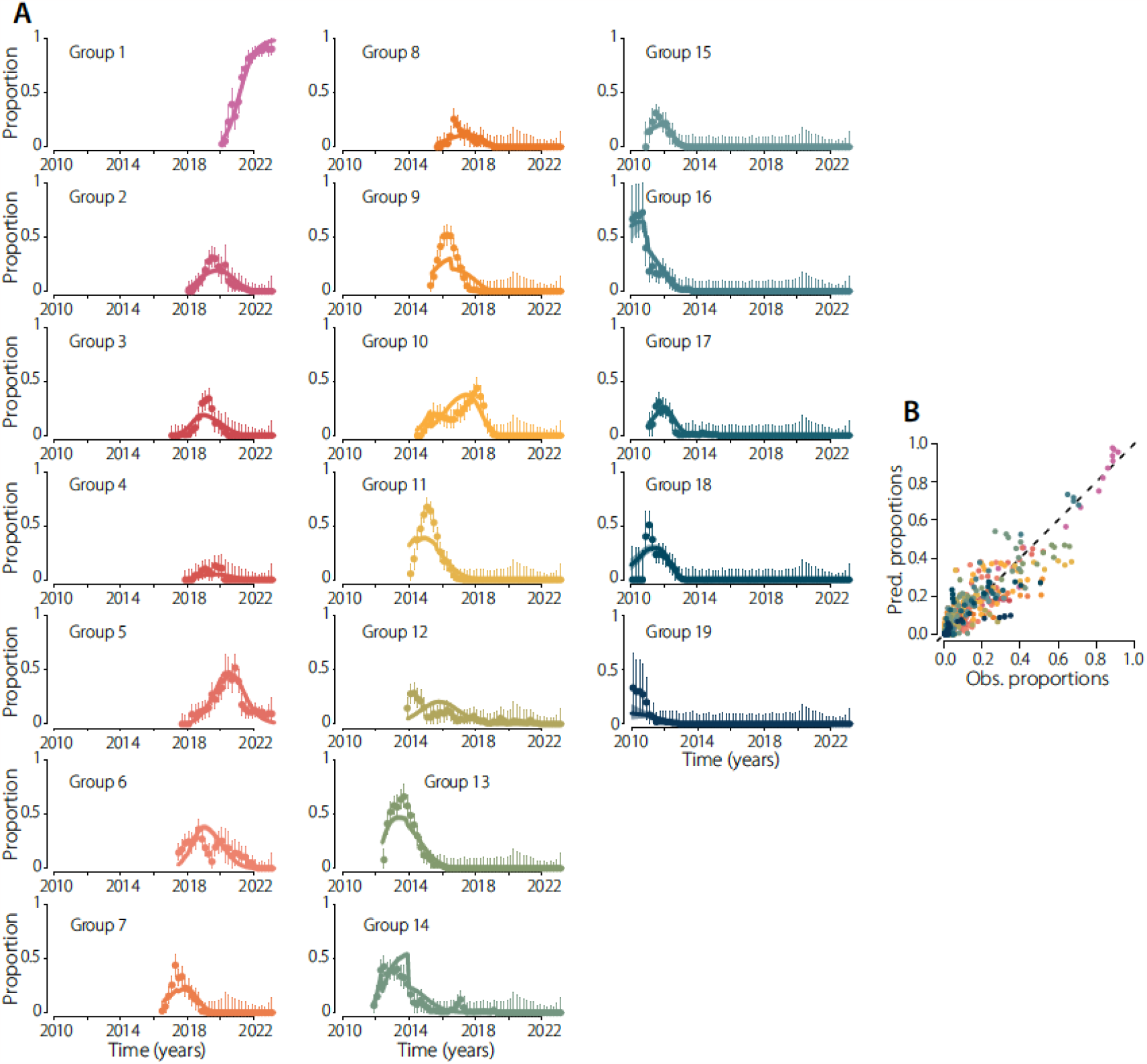
Fitness model fits for all lineages of H3N2. **(A)** Fits of the proportion of all the H3N2 lineages. Colored dots represent data, bars denote 95% confidence intervals. Colored lines and shaded areas represent the median and 95% credible interval of the posterior. **(B)** Predicted versus observed proportions.

**Figure S8:**
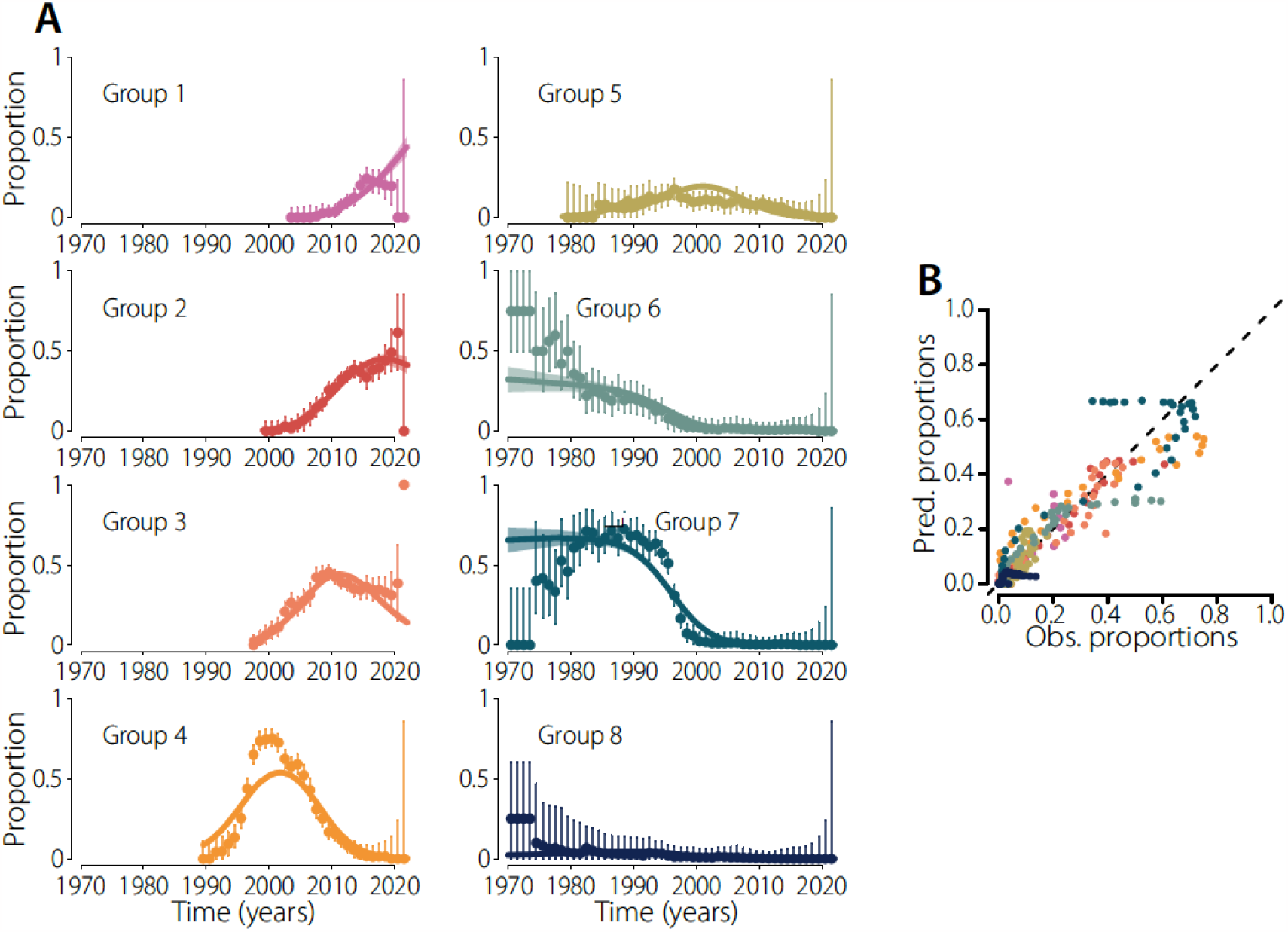
Fitness model fits for all lineages of *B. pertussis*. **(A)** Fits of the proportion of all the *B. pertussis* lineages. Colored dots represent data, bars denote 95% confidence intervals. Colored lines and shaded areas represent the median and 95% credible interval of the posterior. **(B)** Predicted versus observed proportions.

**Figure S9:**
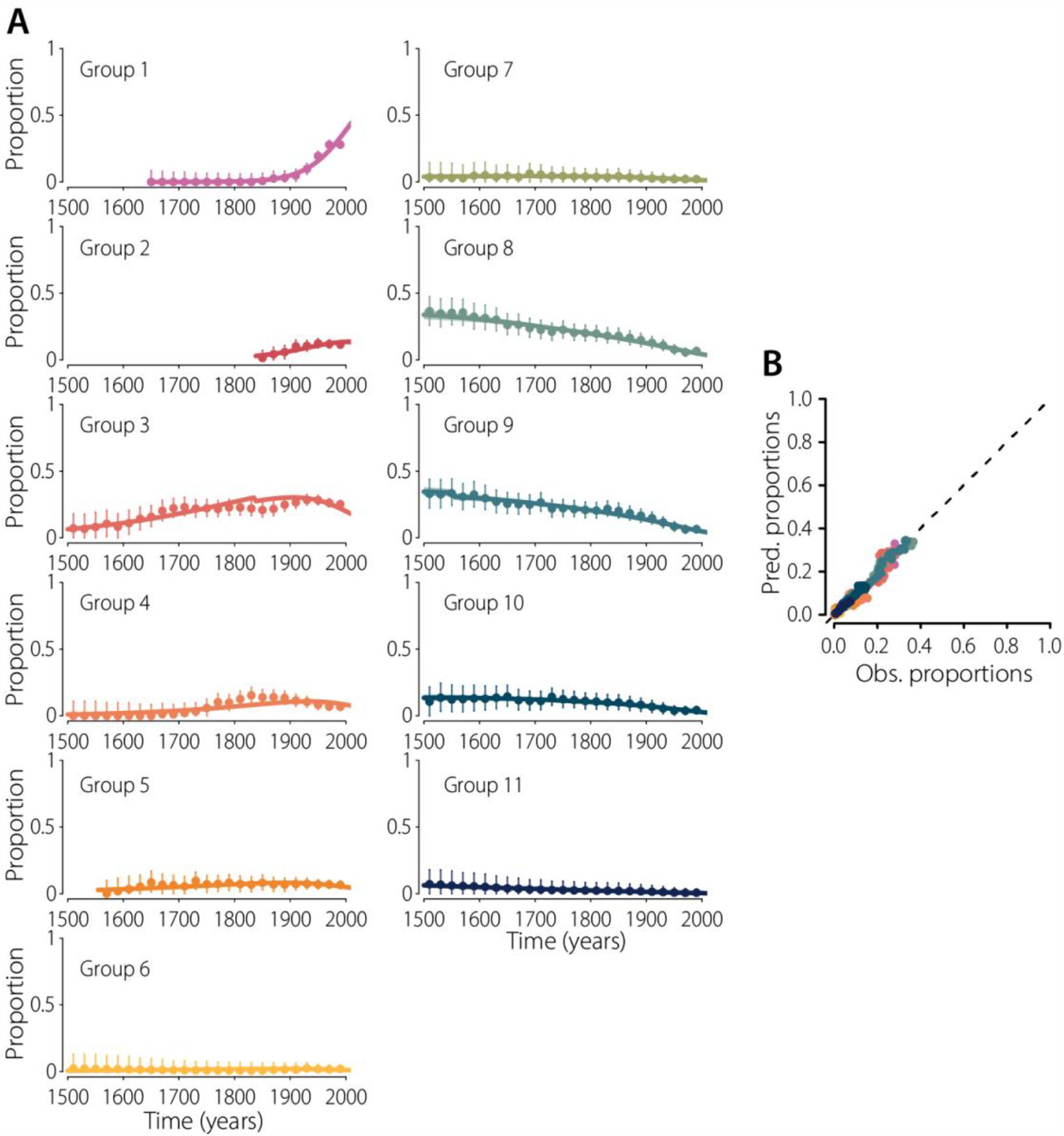
Fitness model fits for all lineages of *M. tuberculosis*. **(A)** Fits of the proportion of all the *M. tuberculosis* lineages. Colored dots represent data, bars denote 95% confidence intervals. Colored lines and shaded areas represent the median and 95% credible interval of the posterior. **(B)** Predicted versus observed proportions.

**Figure S10:**
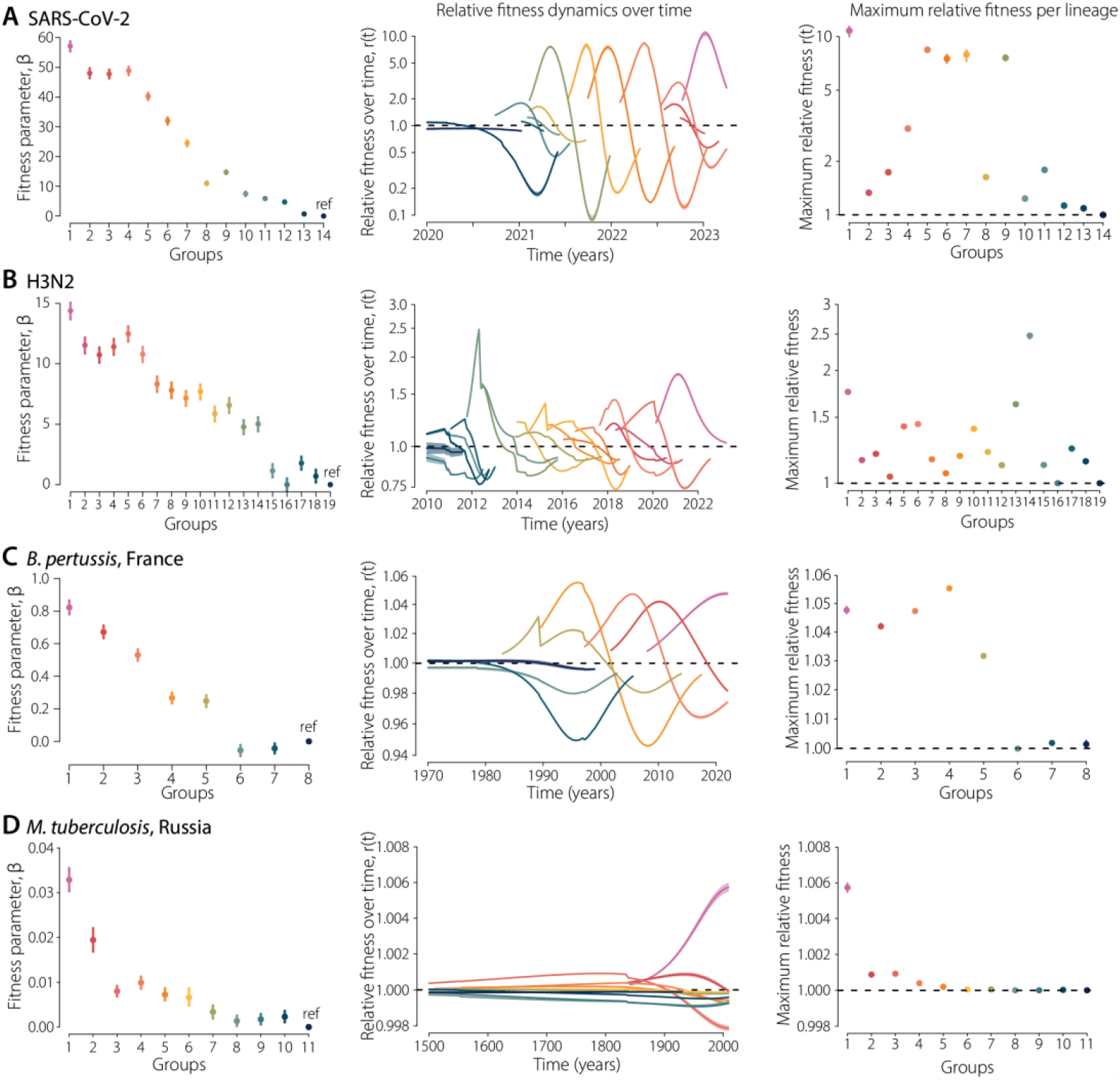
Fitness estimates for all pathogen lineages. For each pathogen we present the estimated fitness of each of their lineages. From left to right: Fitness parameter *β* for each lineage; Relative fitness dynamics overtime *r(t)*; maximum relative fitness per lineage. Dots represent median estimates for each lineage, bars denote 95% credible interval of the posterior. Lines and shaded areas represent the median and 95% credible interval of the posterior. Colors represent the different lineages identified for each pathogen.

**Figure S11:**
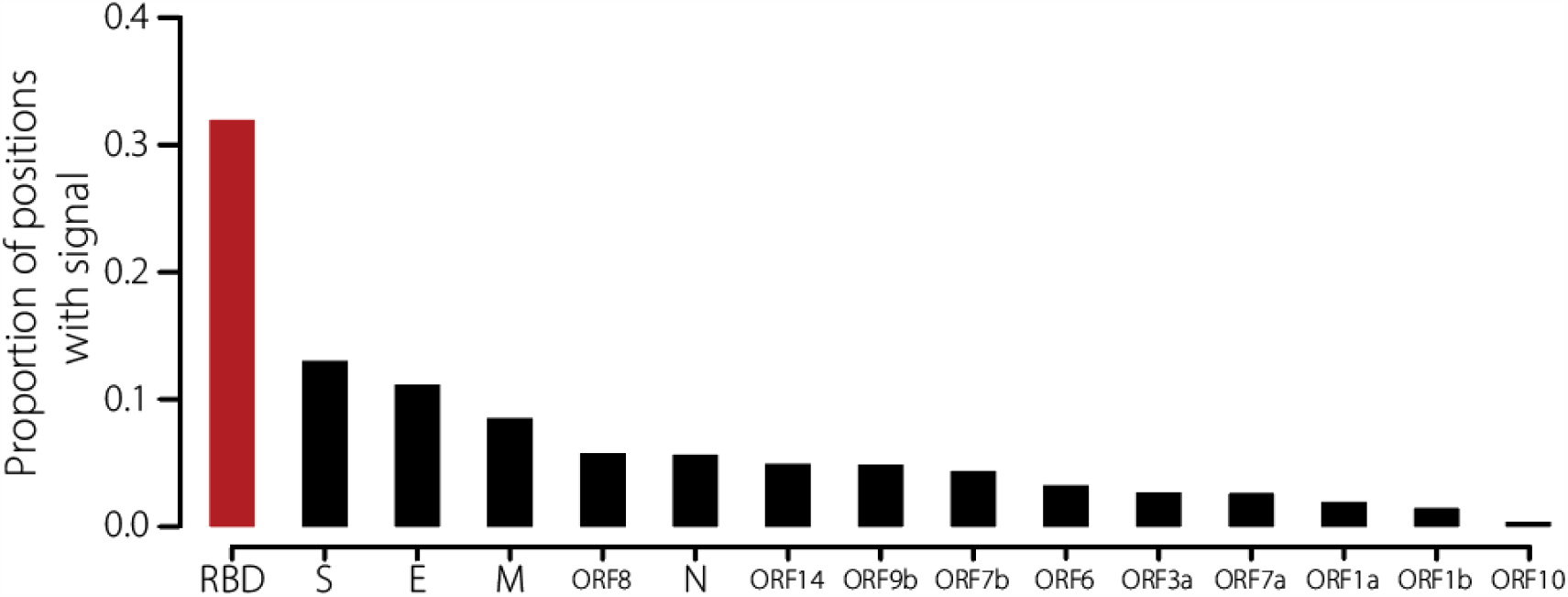
Proportion of mutations that are defining the lineages of SARS-CoV-2 worldwide, by ORFs. Additionally,to Figure 4E, we plot the proportion of amino acid substitutions that are lineage-defining within SARS-CoV-2 ORFs, and the Receptor Binding Domain (RBD).

**Figure S12:**
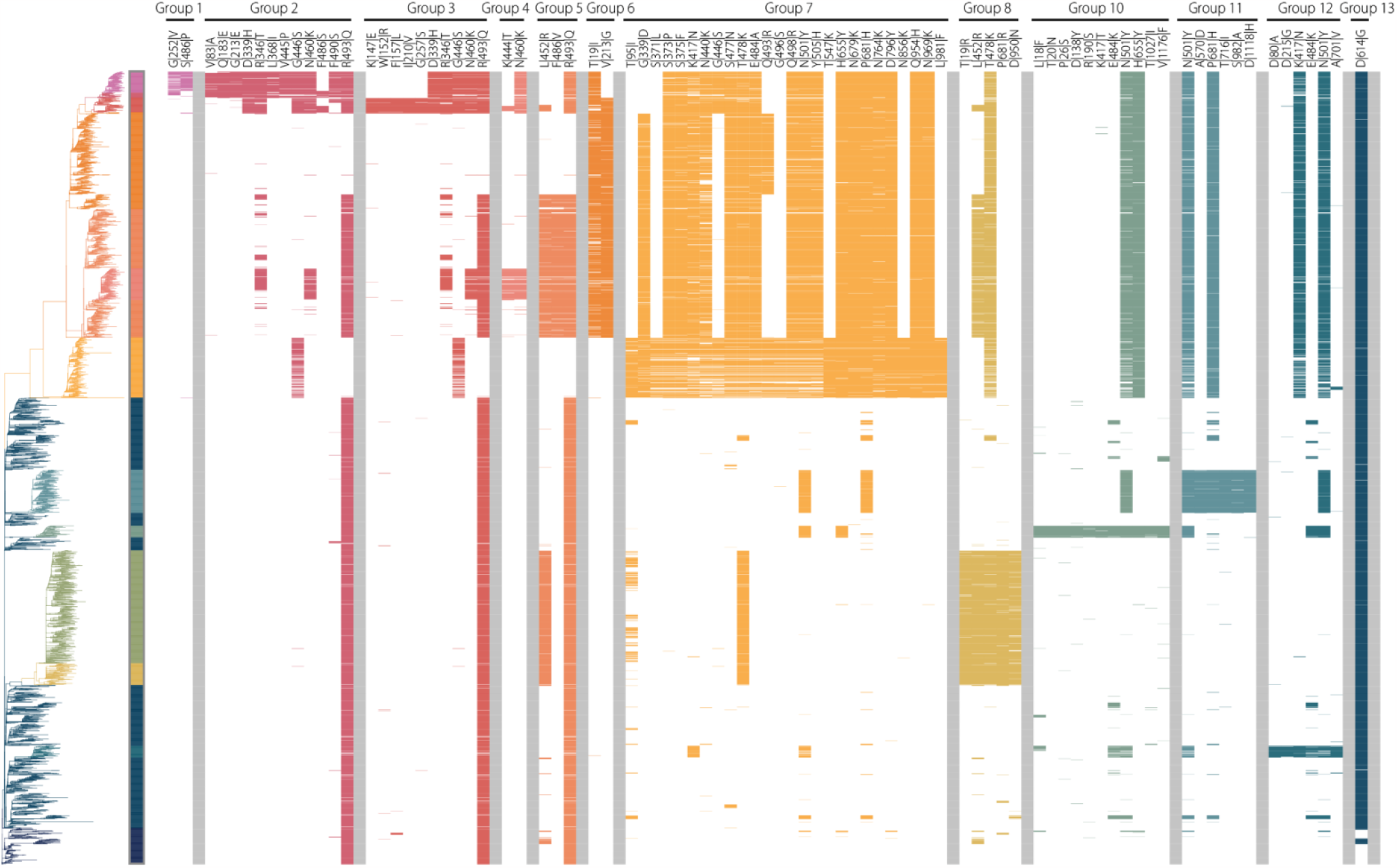
Phylogenetic tree and mutations in the spike protein that are defining lineages in the global SARS-CoV-2 dataset. We present the H3N2 time-resolved tree (left), together with the mutations that we found to be defining its lineages (right). Colors represent the different lineages. Each column on the right displays one mutation, with its name at the top. Colors denote isolates that are carrying the labeled mutation, white denotes the absence of that mutation (although isolates could have other mutations at this position). Some mutations (e.g., T478K or N501Y) are defining multiple lineages and are therefore plotted twice. The list of lineage-defining mutations can be found in Data File S5.

**Figure S13:**
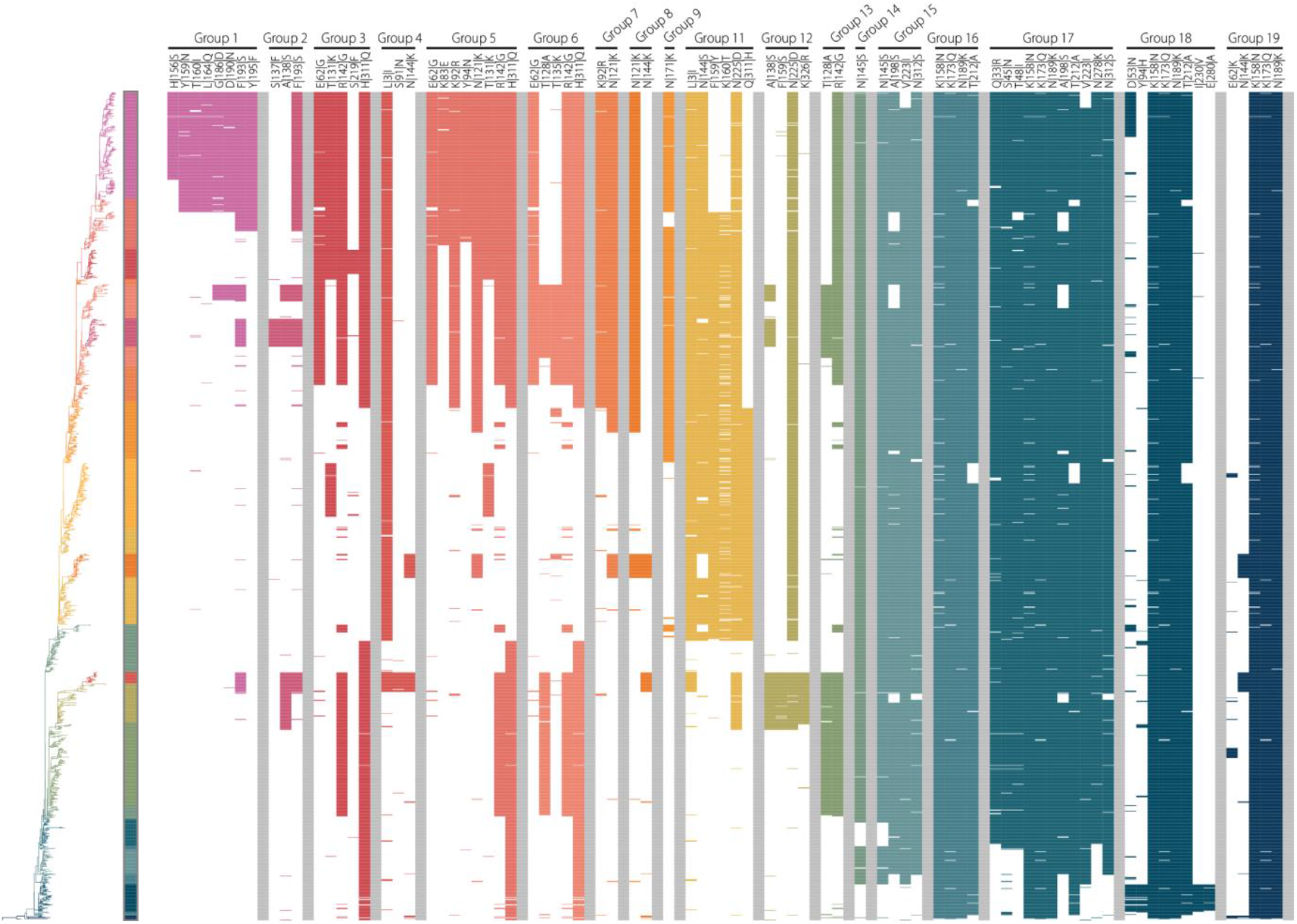
Phylogenetic tree and mutations in the HA1 subunit that are defining lineages in the global H3N2 dataset. We present the H3N2 time-resolved tree (left), together with the mutations that we found to be defining its lineages (right). Colors represent the different lineages. Each column on the right displays one mutation, with its name at the top. Colors denote isolates that are carrying the labeled mutation, white denotes the absence of that mutation (although isolates could have other mutations at this position). Some mutations (e.g., N144K or F193S) are defining multiple lineages and are therefore plotted twice. The list of lineage-defining mutations can be found in Data File S6.

**Figure S14:**
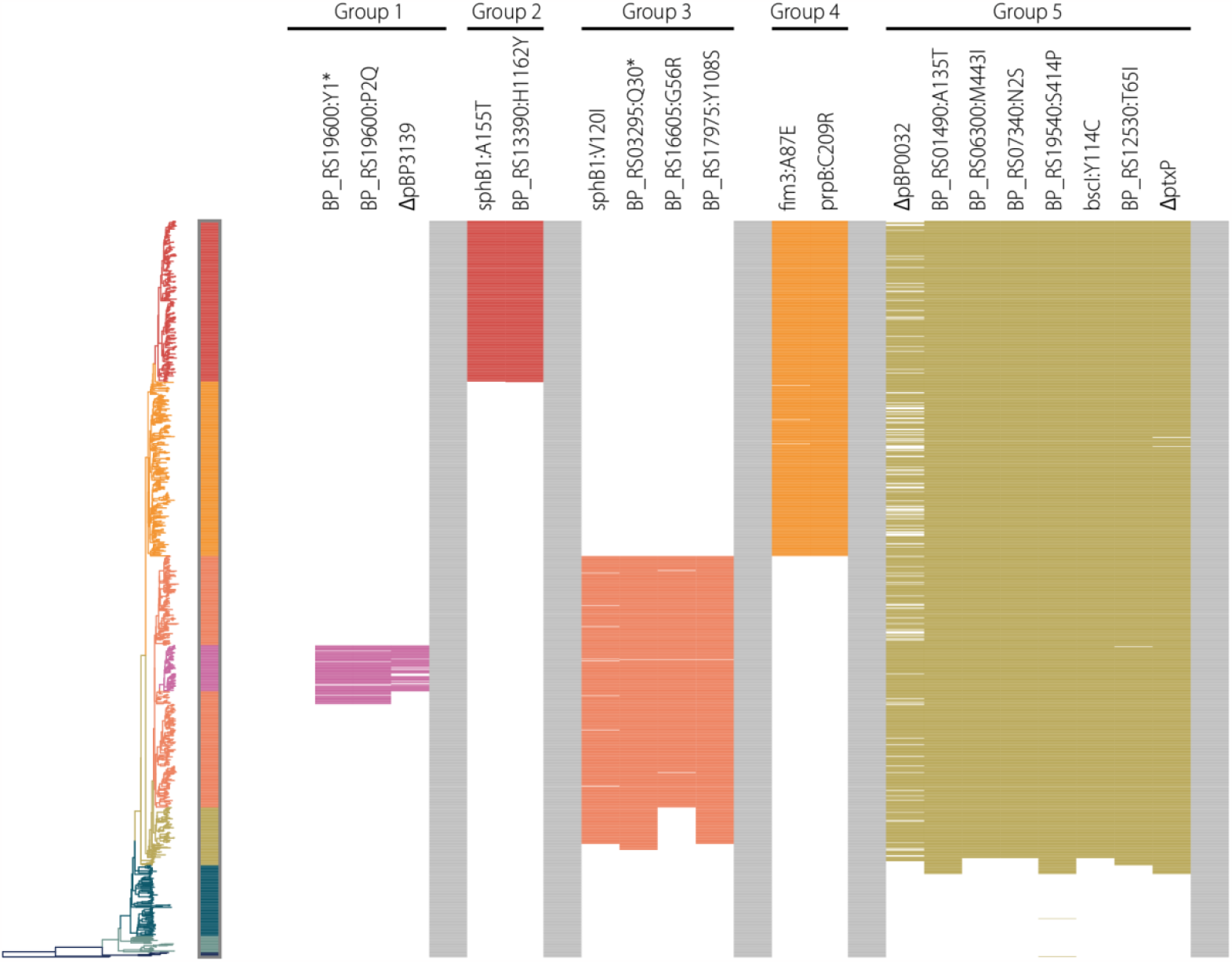
Phylogenetic tree and mutations defining lineages in the *B. pertussis* dataset from in France. We present the *B. pertussis* time-resolved tree (left), together with the mutations that we found to be defining its lineages (right). Colors represent the different lineages. Each column on the right displays one mutation, with its name at the top. Colors denote isolates that are carrying the labeled mutation, white denotes the absence of that mutation (although isolates could have other mutations at this position). The list of lineage-defining mutations can be found in Data File S7.

**Figure S15:**
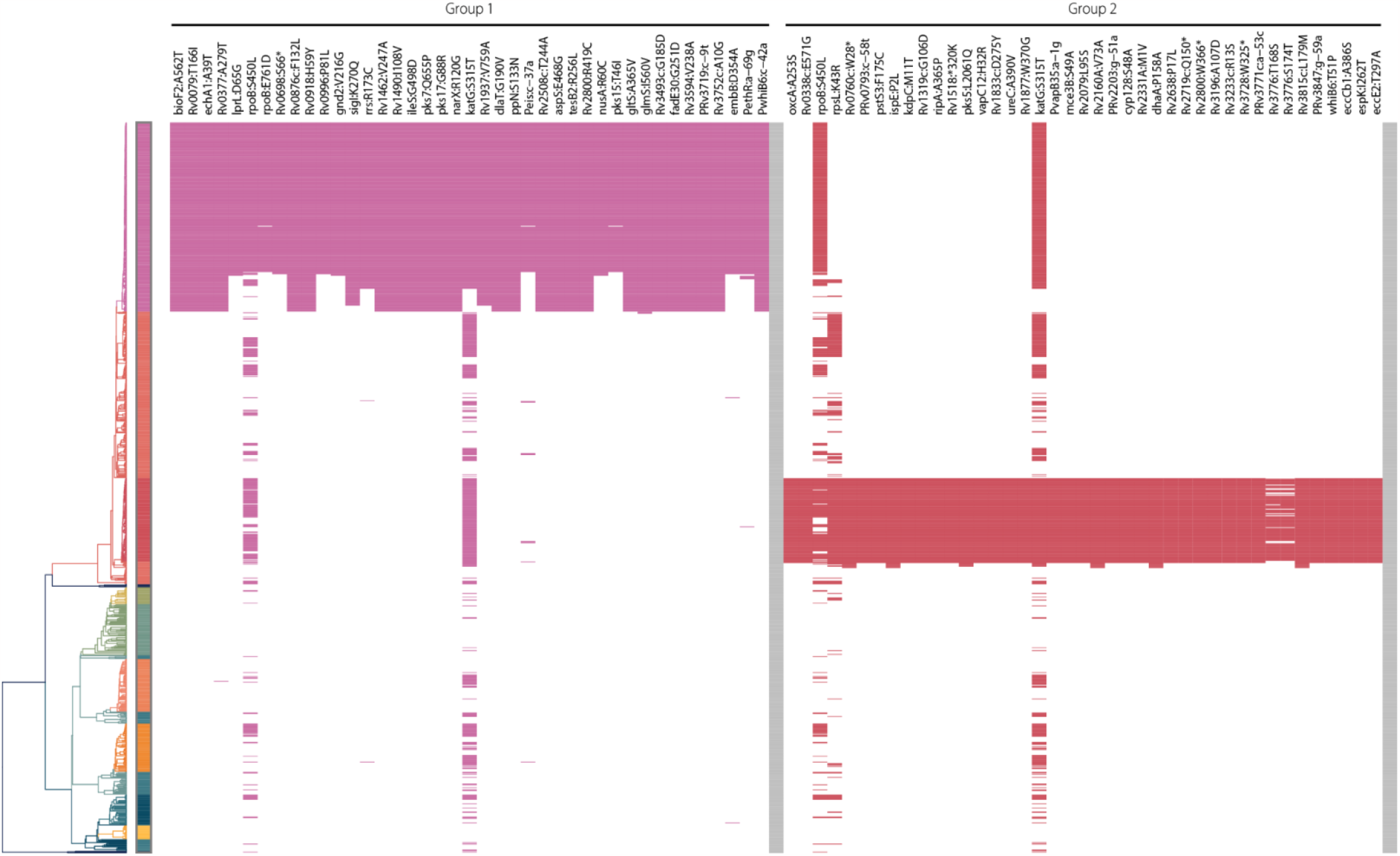
Phylogenetic tree and mutations defining lineages 1 and 2 in the *M. tuberculosis* dataset from in Samara, Russia. We present the *M. tuberculosis* time-resolved tree (left), together with the mutations that we found to be defining the lineages 1 and 2 (right). Colors represent the different lineages. Each column on the right displays one mutation, with its name at the top. Colors denote isolates that are carrying the labeled mutation, white denotes the absence of that mutation (although isolates could have other mutations at this position). Some mutations (e.g., rpoB:S450L or katG:S315T) are defining both lineages and are therefore plotted twice. The list of lineage-defining mutations can be found in Data File S8.

**Figure S16:**
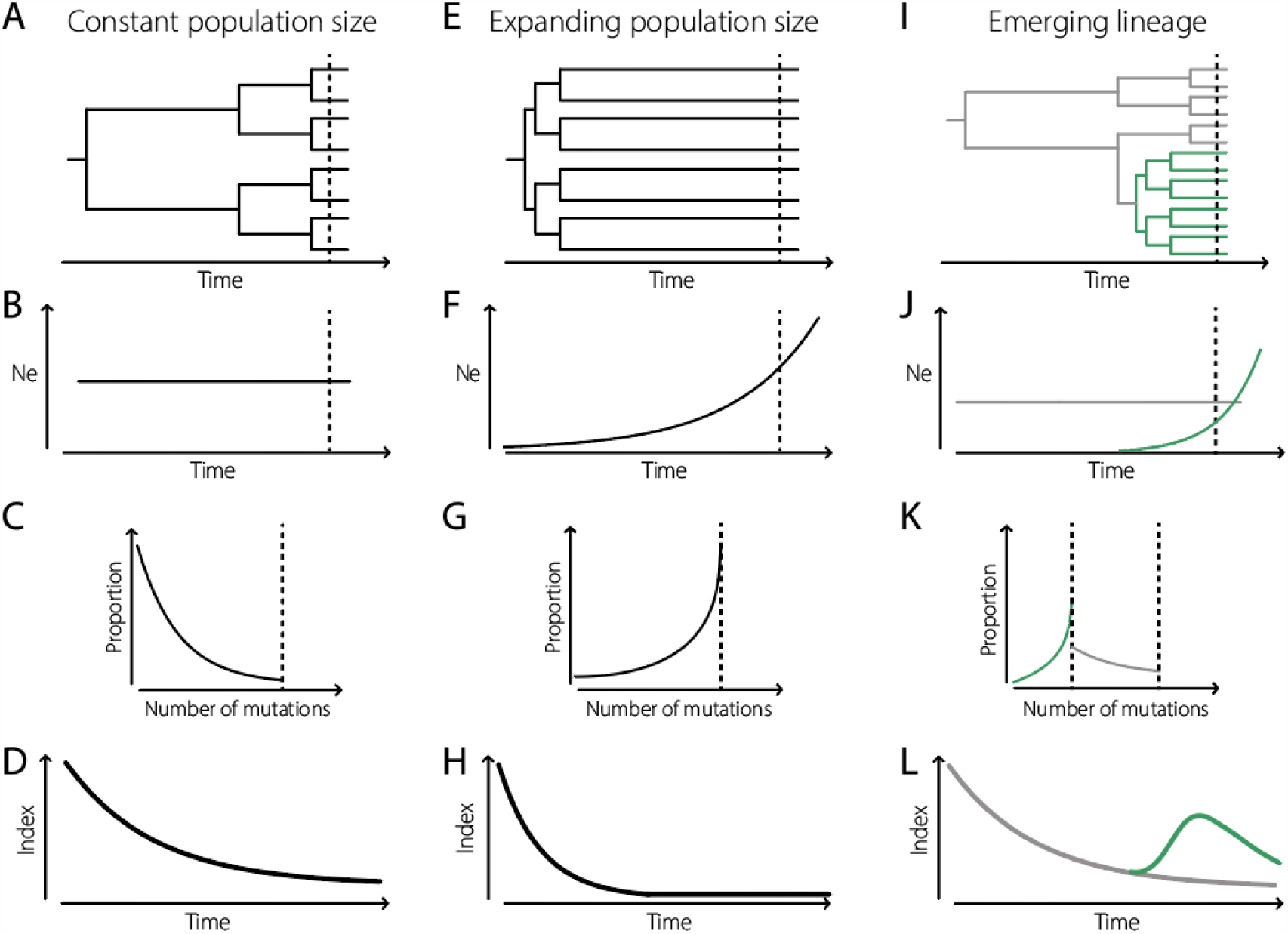
Population history, pairwise distance distribution and index dynamics. **(A-D)** Constant effective population size. **(E-H)** Exponential population size. (A and B are inspired by Volz and colleagues, 2013(*60*)) **(I-L)** Case of an emerging, exponentially growing, lineage in a population of constant effective size.

**Figure S17:**
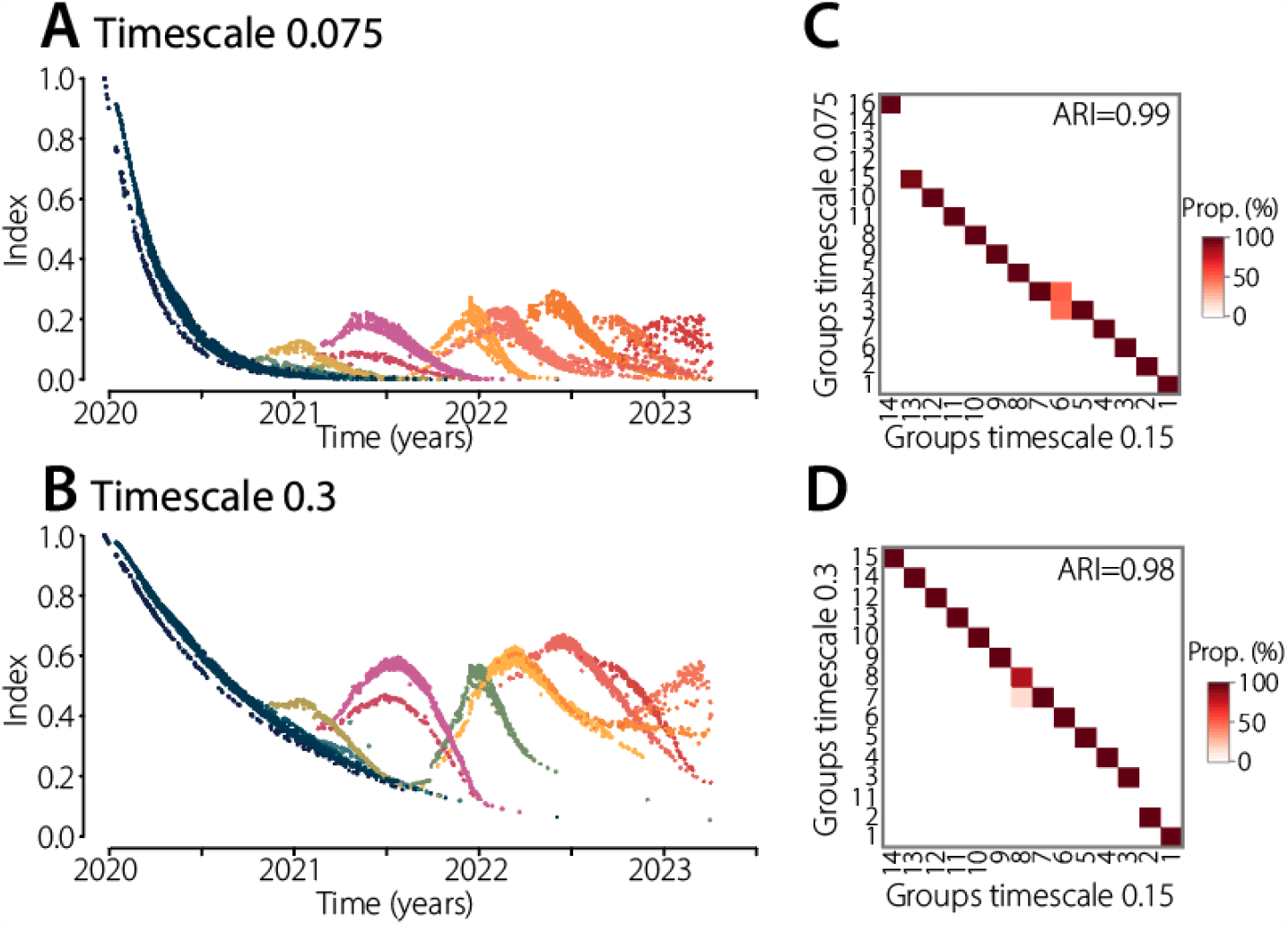
Robustness of the framework to the choice of timescale. We show our framework is robust to the choice of the timescale (governing the weight distribution used in the index computation). **(A-B)** Index dynamics computed on the global SARS-CoV-2 tree, with either a timescale of 0.075 (A) or 0.3 (B). The timescale used in the main analysis is 0.15. A smaller timescale focused more on recent population dynamics; a larger timescale focused more on the past evolution. Colors represent the lineages identified with our algorithm on those dynamics. **(C-D)** We compare the lineages identified with those timescales (y-axis) to the lineages presented throughout this study, with a timescale of 0.15 (x-axis). Darker colors represent more agreement between both classifications. Overall, we find minimal differences in the lineages detected.

**Figure S18:**
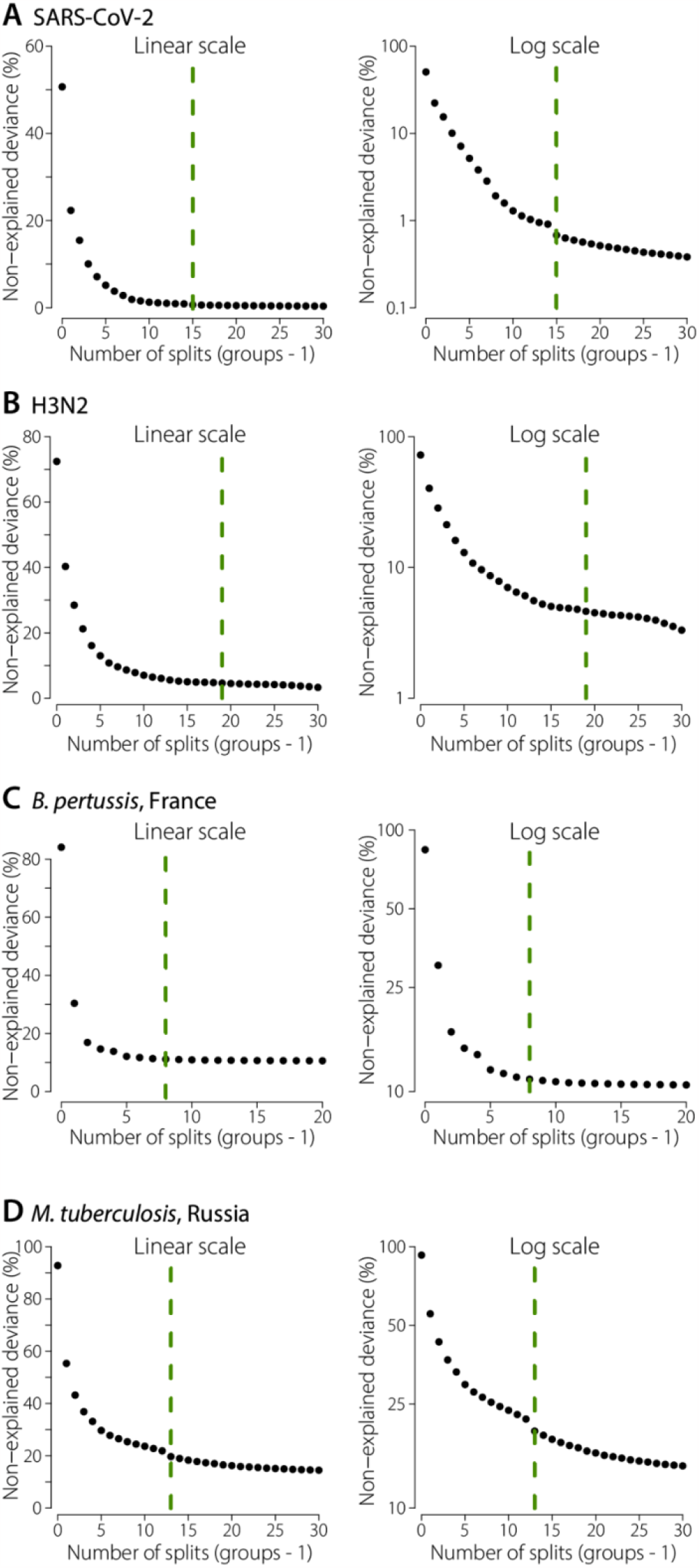
Non-explained deviance as a function of the number of groups in the lineage detection algorithm. For each pathogen, we plot the proportion of non-explained deviance by the models with different numbers of groups. Dashed lines represent the number of groups chosen. We plot the proportion both on a linear scale (left) and log scale (right). The log scale enables a more precise appreciation of the number of groups at which the deviance explained does not increase substantially anymore.

**Figure S19:**
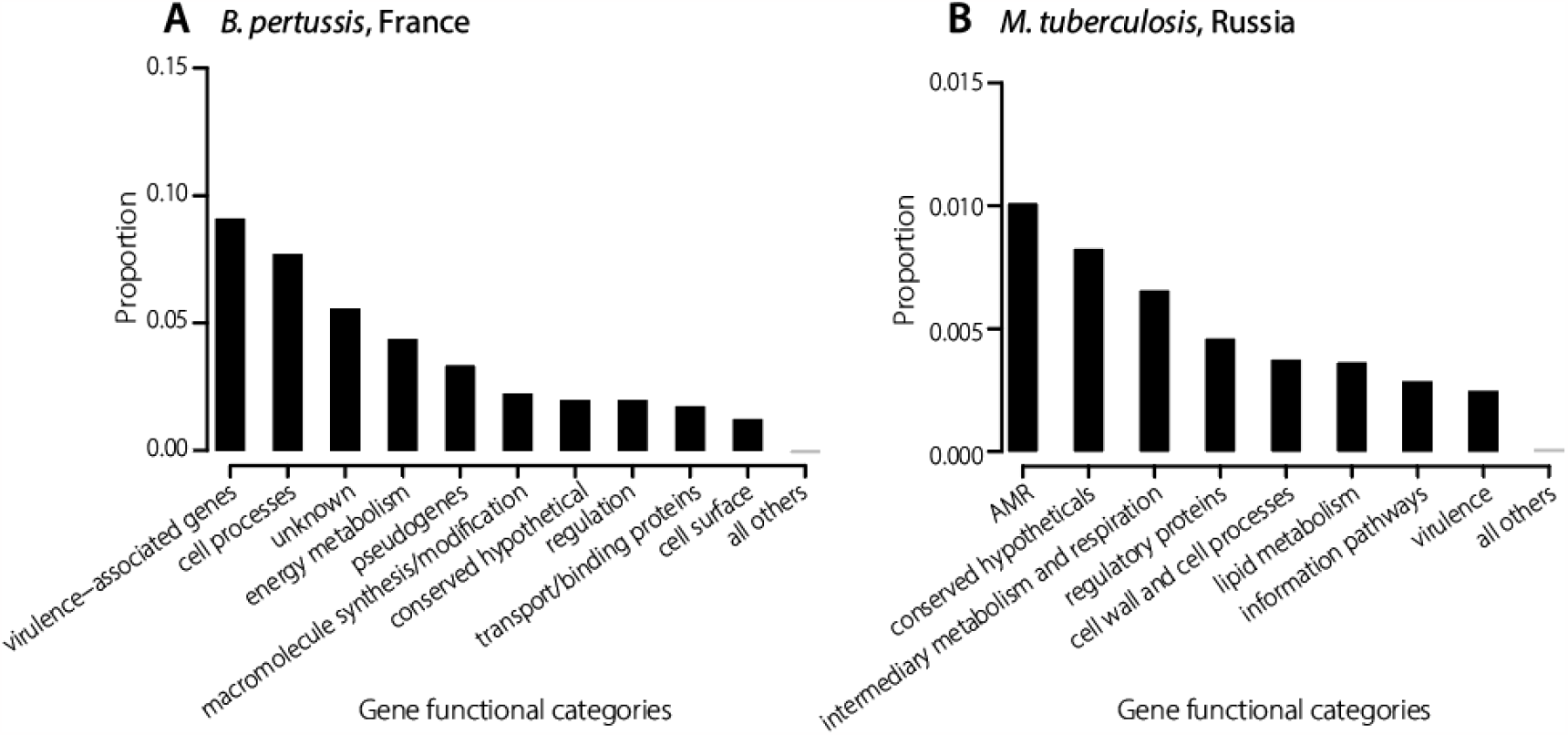
Proportion of synonymous mutations that are lineage-defining, by gene functional categories, for *B. pertussis* and *M. tuberculosis*. Similarly to Figure 4K-L, we plot the proportion of synonymous mutations that are lineage-defining within each functional category, for (A) *B. pertussis* and (B) *M. tuberculosis (36, 37*). For *M. tuberculosis*, we only considered the most recent lineages 1 and 2. As expected, we find no statistical differences, as opposed to Figure 4K-L.

**Figure S20:**
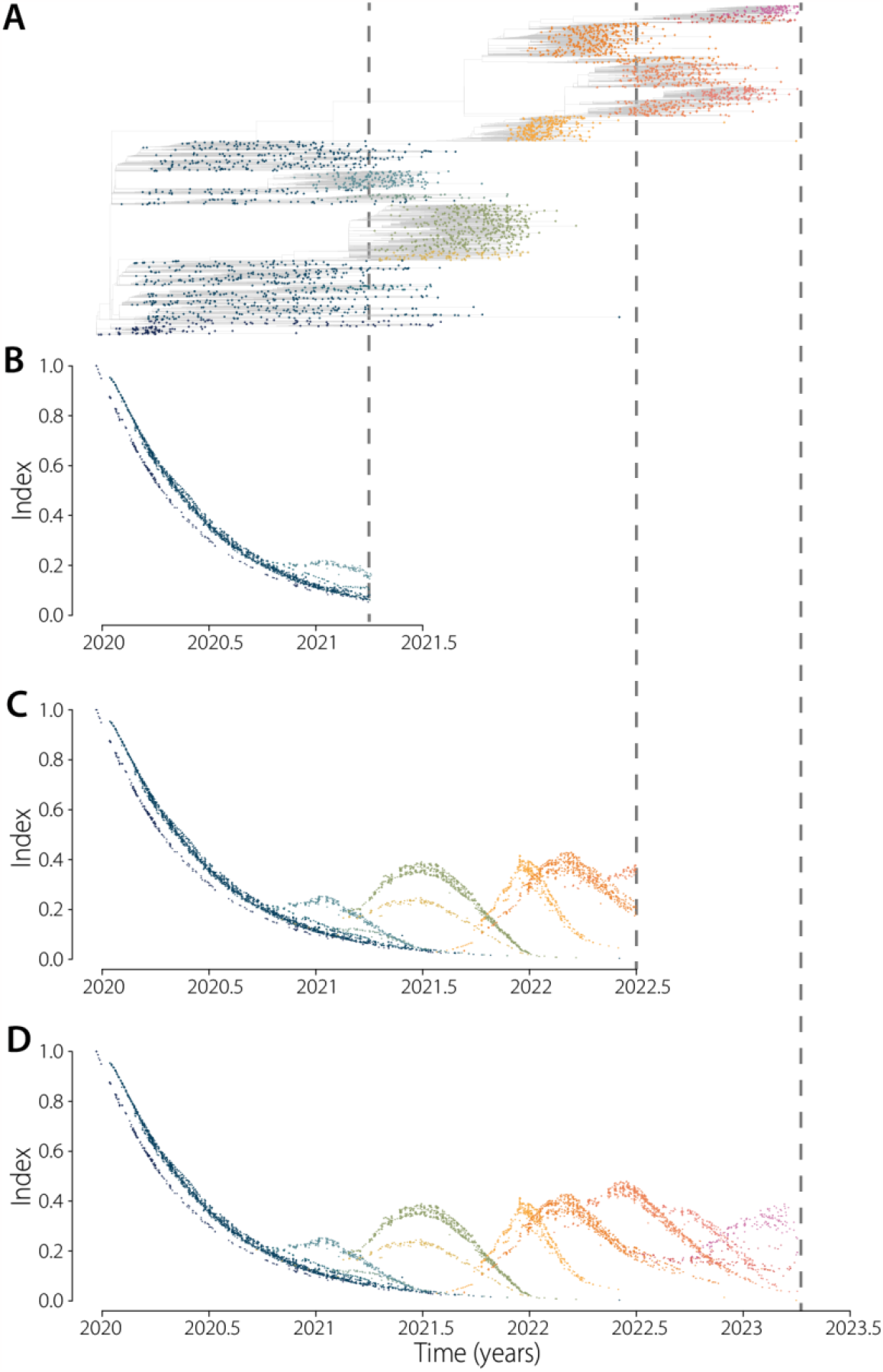
Example of index dynamics on time censored global SARS-CoV-2 datasets. **(A)** Global SARS-CoV-2 time-resolved phylogenetic tree, same as on Figures 1-2. Dots denote terminal nodes only. **(B-C)** Index computed on censored datasets, on either 2021.26 (B) or 2022.5 (C). **(D)** Uncensored index dynamics. When censoring a dataset, we prune all isolates not selected, effectively removing internal nodes and well as terminal nodes. This explains the slightly different dynamics observed near the censoring date. Colors represent the lineages automatically found by our framework (same as Figures 1-2).

**Figure S21:**
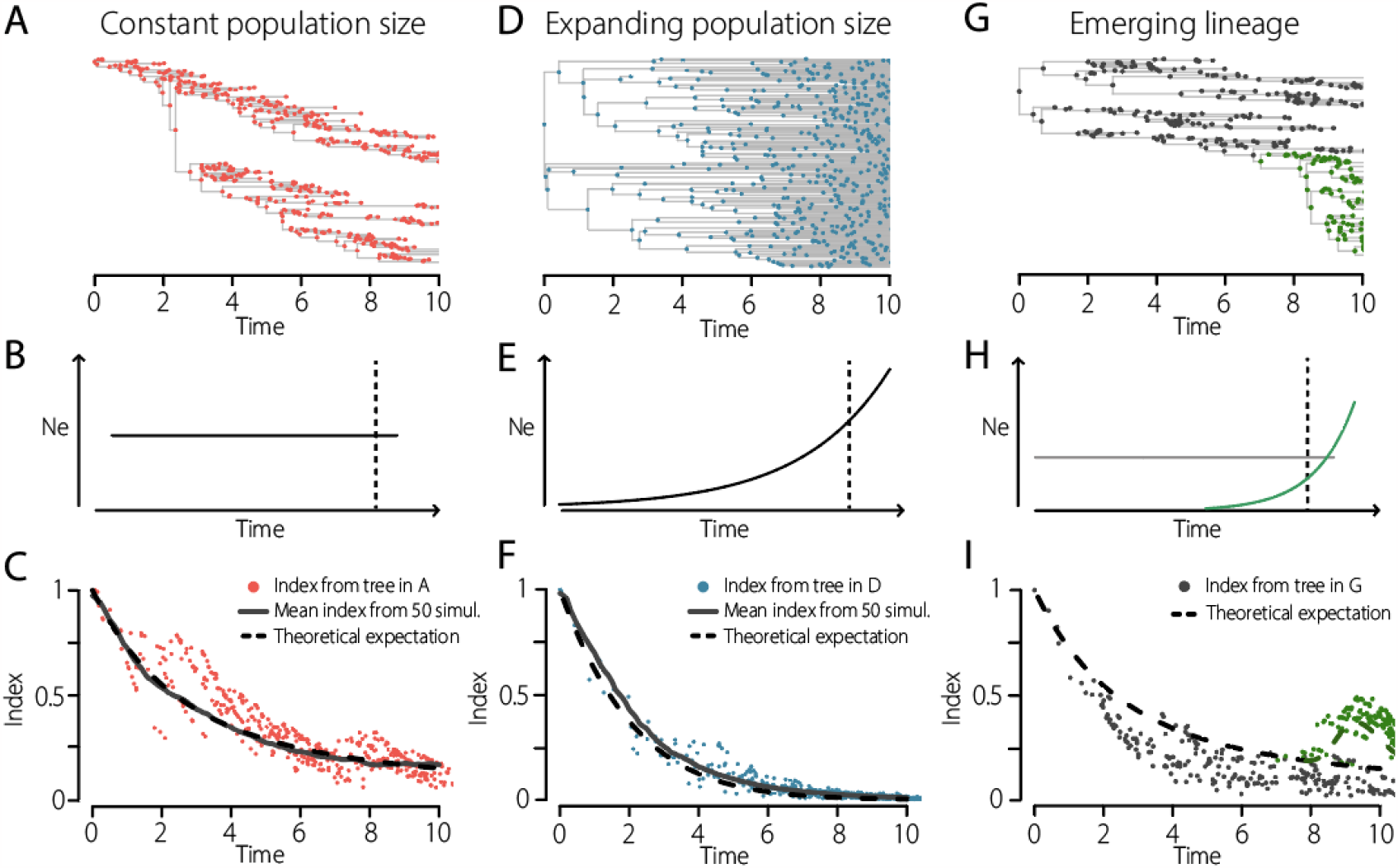
Illustration of the index behavior in different population histories. Similarly to Figure S12, we illustrate here the behavior of the index. In each case, we simulate trees and compute the index on them. **(A-C)** Constant population size. Simulated time-resolved tree, under a birth-death model with equal probability of birth and death, i.e., constant population size on average. (B) Effective population size used in the simulation. (C) Index for through time. **(D-F)** Exponential population size. **(G-I)** Case of an emerging, exponentially growing, lineage in a population of constant effective size. Colors denote each simulation. Dashed lines: expected dynamics given equations in the Methods. Solid lines: mean over the 50 simulations.

**Figure S22:**
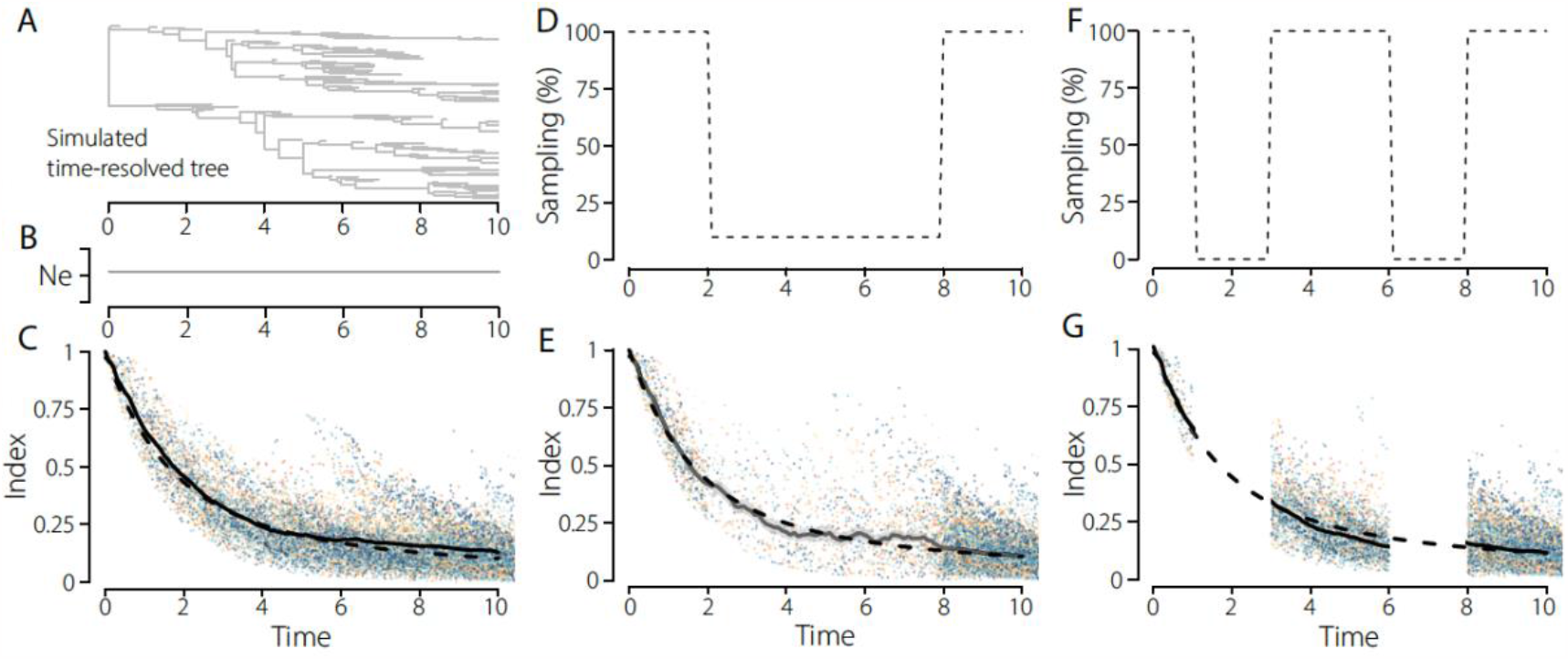
Robustness to sampling schemes, from simulation study. We assess the robustness of the index computation to sampling intensity. **(A-C)** Simulations with no sampling bias. 50 simulations were performed. The tree in A represents one simulation. B represents the effective population size trend: constant. **(D-E)** For each simulation, only 10% of the sequences from year 2-8 were used to compute the index. **(F-G)** No sequences from years 1-3 or 6-8 were used to compute the index. In C, E and G, colors denote each simulation. Dashed lines: expected dynamics given equations in the Methods. Solid lines: mean over the 50 simulations, for the different sampling biases.

